# Hand motor recovery from post-stroke chronic severe hemiplegia with brain-computer interface based neurofeedback exercise: A randomized controlled trial

**DOI:** 10.1101/2024.06.11.24308782

**Authors:** Katsuhiro Mizuno, Masaaki Hayashi, Seitaro Iwama, Daisuke Nishida, Kaoru Honaga, Hiroshi Fuseya, Ryotaro Hirose, Kohei Okuyama, Miho Ogura-Hiramoto, Michiyuki Kawakami, Junichi Ushiba, Meigen Liu

**Affiliations:** Department of Rehabilitation Medicine, Keio University School of Medicine, Tokyo, Japan; Department of Biosciences and Informatics, Faculty of Science and Technology, Keio University, Kanagawa, Japan

## Abstract

**Background:** Post-stroke hemiplegia is a common neurological condition with a significant impact on the daily lives of patients, their families, and society. Since motor imagery and execution are supposed to share common neural networks, repeated motor imagery has been used as a common exercise for motor recovery. However, recent meta-analysis has reported inconsistent results. In the present study, we investigated whether real-time feedback of cortical activities estimated from scalp electroencephalogram (EEG) could enhance the efficacy of motor imagery-based rehabilitation in post-stroke hemiplegic patients.

**Methods:** Forty post-stroke chronic hemiplegic patients received 10 days of 40-minute mental practice with motor imagery of the paralyzed hand in addition to 40-minute standard occupational therapy. Patients were randomly allocated to REAL and CTRL groups. In the REAL group, the EEG sensorimotor rhythm recorded over the ipsilesional sensorimotor area were fed back during motor imagery though combination of visual cursor movement, neuromuscular electrical stimulation, and robotic movement support. The patients in the CTRL group wore the same EEG headset as the REAL group, but no feedback was provided. The primary outcome was the upper-extremity subscales of the Fugl-Meyer assessment (FMA). Primary outcomes were tested after 28 days of intervention using analysis of covariance models. This trial is registered with UMIN Clinical Trial Registry, UMIN000026372.

**Findings:** FMA scores showed a significant main effect of time (p = 0·027) with an ANCOVA model considering baseline but not a significant between-group difference. However a significant interaction between time and group (p = 0•037) and greater functional gain in the REAL group (p < 0•001) were found with another model considering patient heterogeneity. The improvement was accompanied with the significant changes in the modulation index of finger extensor muscles in the affected hand in the REAL group. Moreover, imagery-related cortical activation in the sensorimotor area was significantly greater in the REAL group than in the CTRL group. No moderate or serious adverse events were confirmed in both groups.

**Interpretation:** EEG-based brain-computer interface neurofeedback may enhance the efficacy of mental practice with motor imagery and augment motor recovery in poststroke patients with chronic and severe hemiplegia.

**Funding:** Sponsored by AMED under Grant Number JP18hk0102032 and JSPS KAKENHI Grant Number 20H05923.

## Introduction

Every year, 15 million people worldwide suffer a stroke.^1^ Only 25% of stroke survivors recover their motor function^2^, and no further recovery is expected after 6 months from the onset of stroke.^3^ Thus, five million people per year are left permanently disabled.^4^ Disablement results in a loss of independence in activities of daily living (ADL), reduces quality of life (QOL) and increases caregiver burden.

Post-stroke motor impairment is often persistent, since an endogenous process of functional brain remodeling involving regions outside the lesion site halts when it reaches a chronic stage.^5^ Many interventional approaches, such as intensive use of the affected limb combined with restraint of the unaffected limb, mental practice with motor imagery, robotic movement assistance, or neuromodulation through transcranial or peripheral electromagnetic stimulation, have been tested trying to resume neural reorganization, but therapeutic efficacy is limited for hand motor function among these interventions.^6^

Recently, it is found that repeated hand motor imagery in the paretic side, accompanied with exogenously induced somatosensation through Brain-Computer Interface (BCI), has a great potential to facilitate the resumption and shaping of the neural remodeling process for functional recovery of paretic hand movements.^7,8^ BCI for this purpose often use scalp electroencephalogram (EEG) to detect event-related desynchronization (ERD) of ipsilesional sensorimotor Rolandic brain oscillations (sensorimotor rhythm [SMR]), and translate it to the cortical excitability.^7^ BCI gives visual feedback of this ongoing cortical excitability estimated from SMR-ERD magnitude, and triggers functional electrical stimulation (FES)/robotic assistance to the paretic extremity when the excitability exceeds the pre-determined threshold. Through this framework, patients learn to control brain oscillatory activity and related excitability involving regions outside the lesion site, while attempting paretic hand movement. Clinical studies have revealed resumed functional reorganization in the ipsilesional sensorimotor network in the brain.^9^

Nowadays, thirteen randomized control trials have tested clinical efficacy of BCI-based therapy over the randomly actuated FES and robotic assistance. A systematic review (incorporating 235 patients) showed a moderate-to-large effect size in clinical motor improvement, beyond minimally clinical important difference.^8^ It is therefore hypothesized that superior hand motor improvement in severely brain damaged post-stroke patients could be induced if a close contingent connection between the neural correlate of the sensorimotor cortex excitability and the consequent feedback of the movement (visual and proprioceptive) is established via a BCI. However, the clinical effectiveness, that can be concluded by the comparison with the standard therapy for the severe post-stroke hemiplegia remains unclear.

Here, the present clinical trial challenged to test the clinical effectiveness of BCI-based treatment for post-stroke motor recovery (1) to see functional motor improvement in the post-stroke chronic stage (> 90 days) with the severest motor deficits, in which no further recovery is expected, and also, (2) to see functional motor improvement in hand function, for which no accessible and effective treatment has been developed. In the control group, we employed simple motor imagery training with the same dose as the BCI-neurofeedback intervention group. Behavioral physiotherapy was applied for both groups to see the adjuvant effects of BCI-neurofeedback intervention compared with the control. We speculated that learning to control oscillatory brain activity through this BCI approach constitutes the necessary therapeutic ingredient for hand motor recovery in the above-mentioned patient group, and that physiotherapy allows generalization of relearned motor skills to meaningful real-life activities.

## Methods

### Study design

We performed this investigator initiated multi-center, interventional, parallel-group, assessor-blinded, randomized (1:1), trial of BCI versus standard MI intervention, in addition to a 40-minute standard occupational therapy, for patients with post-stroke hemiplegia receiving treatment in 4 hospitals in Japan. This study was conducted in accordance with the guideline of Good Clinical Practice. The study protocol has been published.^10^ Ethics approval was granted by the Institutional Review Board of Keio University Hospital, Japan (D16-03, 2022-1092). The collection and management of clinical data were handled using Electronic Data Capture by the Data Management Unit of the Clinical Research Support Division at Keio University Hospital’s Center for Clinical Research Promotion and by A2 Healthcare Corporation. Monitoring, safety information support, and audits were conducted by CMIC Co., Ltd.

### Participants

Prospective participants were recruited from outpatients of the rehabilitation department in main hospital (Keio University Hospital), followed by three affiliated hospitals (Saiseikai Kanagawa-ken Hospital, Tokyo Metropolitan Rehabilitation Hospital, and Tokyo Bay Rehabilitation Hospital). The recruiting physiatrists screened the participants for eligibility.

The total sample size in this trial was set at 40 patients (20 per group). The SD for the change is also assumed to be 4.0. Under these assumptions, 17 patients per group results in 80% power for testing the between-group difference in means with a two-sided 5% alpha. Considering 15% exclusions from the analysis, the target sample size in this trial was set at 20 patients per group (total of 40 patients).

The inclusion criteria were (1) time from stroke onset to be more than 90 days; (2) first ever stroke patients with upper extremity paresis; (3) no loss of proprioception in paretic fingers (patients capable of detecting a position change after maximum possible motion); (4) ability to raise the paretic hand to the height of the nipple; (5) passive range of motion greater than −10 degrees for metacarpophalangeal joint extension; (6) ability to flex the paretic fingers voluntarily but not to extend them; (7) ability to walk independently in daily life with or without assistive device; (8) ability of the patient to understand and consent to the study protocol; and (9) aged 18 years or older at the time of agreement to participate in this study.

The exclusion criteria were (1) serious medical conditions that would interfere with rehabilitation such as severe heart disease, uncontrolled hypertension, history of pulmonary embolism, acute cardiorespiratory disease or severe pulmonary hypertension within 90 days before enrollment, severe hepatic or renal dysfunction, severe orthopedic impairment, severe cognitive or psychiatric disorder, and other serious medical conditions; (2) use of a pacemaker or of other implanted stimulators; (3) history of seizures within 90 days before enrollment; (4) participation in another clinical trial for regulatory approval within 90 days before enrollment; (5) receiving other special neurorehabilitation techniques for upper extremity paresis such as transcranial magnetic stimulation, therapeutic electrical stimulation, constraint-induced movement therapy (CIMT), and repetitive facilitative exercise within 90 days before enrollment; (6) injection of botulinum toxin or phenol for treatment of upper-limb spasticity within 90 days before enrollment; (7) impossible to record EEG because of skin status or skull deformity; or (8) other critical problems that would hinder participation in the study.

We obtained written informed consent from patients who met all eligibility criteria. The patients then tried the intervention to check the skin status and rule out a skull deformity. If EEG could be recorded, the patient was registered as an eligible and consenting participant. Afterwards, they completed the baseline assessment. Treatment started within 28 days after registration.

Forty poststroke hemiplegic inpatients in total participated in the study; none had participated in any other studies. All participants had normal or corrected-to-normal vision and reported no history of cognitive or psychological disorders. The study was conducted in accordance with the Declaration of Helsinki. The experimental procedure was approved by the Institutional Review Board of Keio University Hospital, Japan (protocol number: KCTR-D008, reference number: D16-03) and originally registered to the national registry system (UMIN000026372; UMIN Clinical Trial Registry (UMIN-CTR)). To perform post-hoc multivariate ANCOVA, the outcomes were analyzed retrospectively under alternative approval of the statistical procedure by the Ethics Committee of the Keio University School of Medicine (Number: 20221092). Written informed consent was obtained from all participants prior to the trial.

### Randomization and masking

Participants were randomly allocated to the REAL and CTRL groups using a computerized block randomization scheme, including prestratification according to each participating hospital. The primary outcome measure was assessed by a blinded evaluator. Other functional measurements were assessed by evaluators who were trained by the organizer of this RCT. The raters were masked to treatment allocation, and they were not involved in the participants’ treatment. The blinded rater assessed FMA and the rater blinding was verified with a yes or no question: Did you rate the assessment score blindly? Participants were not blinded to their own treatment.

### Procedures

The EEG-based BCI neurofeedback system was designed to help mental practice of motor imagery for poststroke severe hemiparetic patients who cannot perform voluntary finger extension, which is a substitute for performing the actual movement exercise. The exercise with this EEG-based BCI neurofeedback system encompasses the following 4 components: standard medical treatments with motor imagery^11^, biofeedback ^12^, neuromuscular electrical stimulation (NMES) to paretic muscles^13^, and robot-aided sensorimotor stimulation^14^. By combining these components, the EEG-based BCI intervention realizes voluntary control of endogenous sensorimotor activities in the ipsilesional hemisphere by actuating the NMES and robotic device contingent on the SMR-ERD magnitude (Figure 2). Detailed information on the apparatus is described in Appendix.

**Figure 1.**
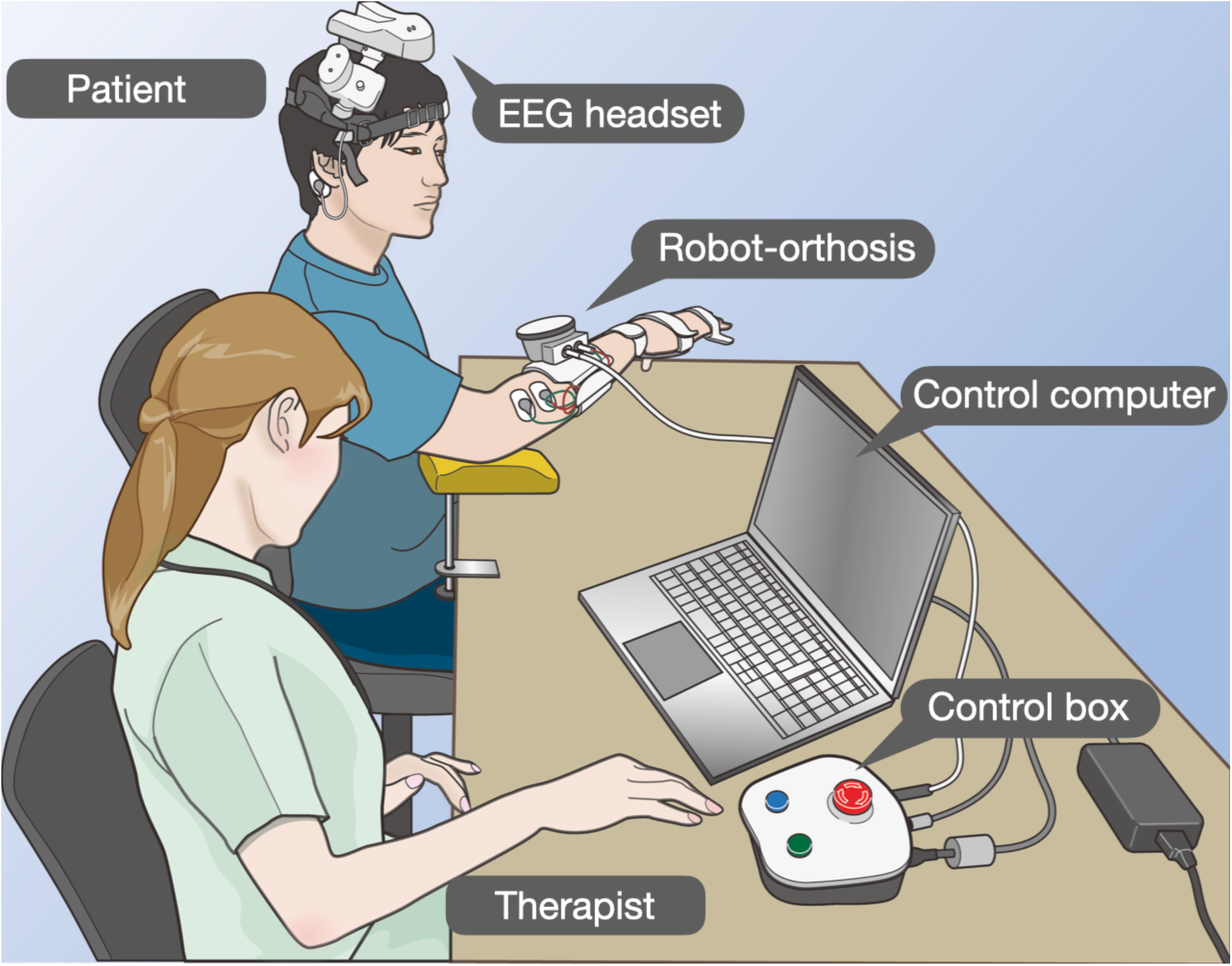
Schematic of EEG-based BCI neurofeedback. The EEG electrodes were placed at C3 (or C4) and 20 mm lateral location that is located closest to the hand region in sensorimotor cortex according to the international 10-20 system. EEG data were transferred to the control computer and processed online. The control computer sent trigger signals to the control box to activate NMES and the robot-orthosis to provide somatosensory feedback.

**Figure 2.**
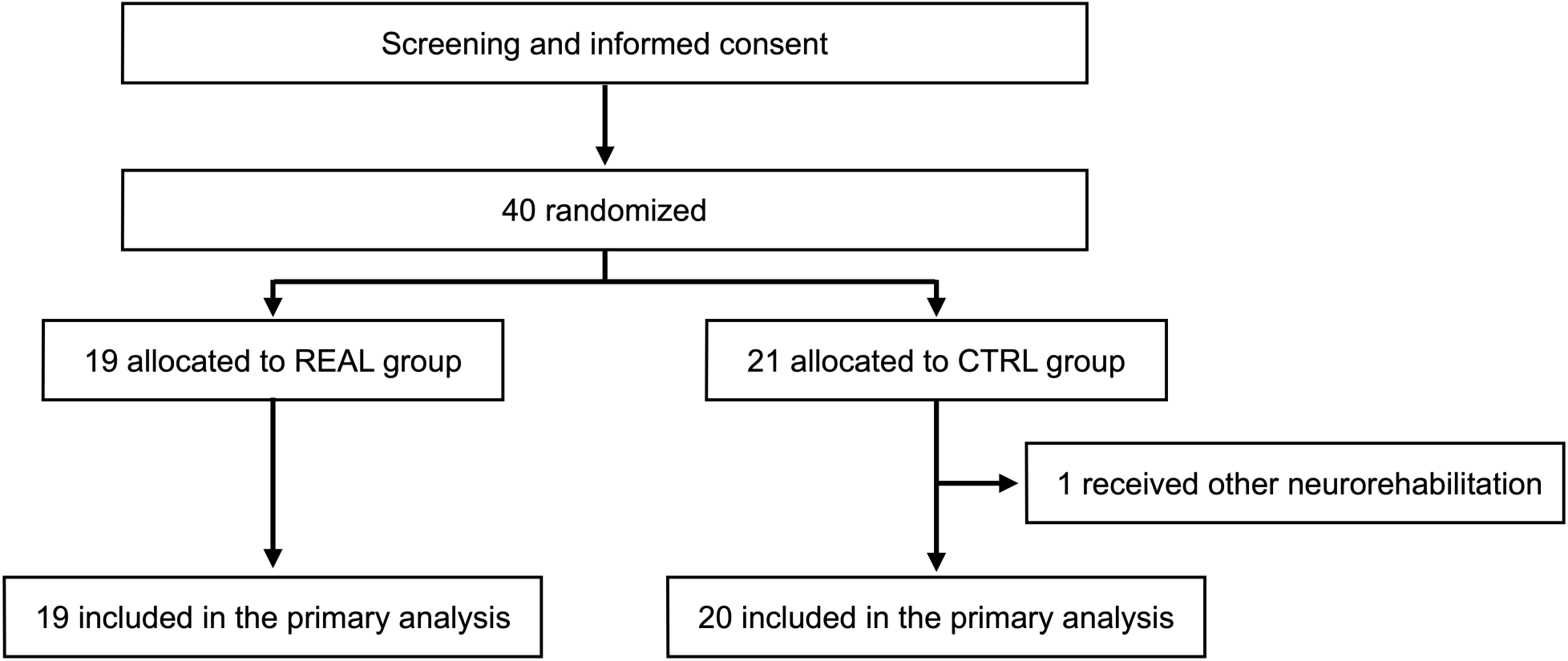
Trial profile. * Receiving other special neurorehabilitation techniques for upper extremity paresis such as transcranial magnetic stimulation, therapeutic electrical stimulation, CIMT, and repetitive facilitative exercise within 90 days before enrollment.

EEG-based BCI neurofeedback exercise was conducted for approximately 40 minutes per session, 1 session per day, and 5 days per week, for 2 weeks. The robot-orthosis and disposable electrodes for NMES were attached to the affected hand to achieve finger extension movement at the metacarpophalangeal and proximal interphalangeal joints. Before starting the exercise routine, the calibration session was performed every day. Five seconds of rest were first given, and a text cue of either “Imagine paretic finger extension” or “Keep relaxing” was then displayed on the top of the computer screen in a random order. The participants performed the given cued task for the next 5 seconds. A total of 20 trials (10 trials per each task) was given, and the EEG classifier based on linear discriminant analysis (LDA) was calibrated using the collected EEG data (Appendix).^15^

During training, patients pinched a peg with the affected hand and then pressed a button to start the exercise sequence. In each sequence, a 5-second resting-state was first given, and a text cue of “imagine paretic finger extension” was then displayed on the top of the computer screen. The participants performed motor imagery of paretic finger extension in the next 5 seconds. The EEG classifier returned either the value +1 (EEG was at the resting condition) or −1 (EEG-SMR-ERD appeared) every 125 ms according to the EEGs, and triggered the robot-orthosis action if a successive 1 second of the class −1 is given in the task period. NMES was applied throughout the time the class was determined to be −1 (Figure S2 in appendix).

Participants in the CTRL group conducted the dose-matched motor imagery exercise for 40 minutes for 2 weeks, resembling the EEG-BCI based neurofeedback exercise in the REAL group.^16^ Participants wore the same EEG headset as those in the REAL group, but they did not wear the hand orthosis. They were instructed to rest for 5 seconds and then to imagine extending their affected fingers for the next 5 seconds in the same manner, as those in the EEG-BCI based neurofeedback exercise in the REAL group. EEG was recorded in the same way as in the REAL group, but no feedback about EEG was given to the participants. Neither NMES nor robot-orthosis action was given during imagery.

In addition to one of the two types of intervention, patients in both groups received occupational therapy. The therapy consisted of gentle stretching exercises, active muscle re-education exercises, and introduction to bimanual activities in their daily lives.

### Outcomes

The primary efficacy outcome was the change in the Fugl-Meyer Assessment (FMA) score between baseline and 28 days after the study treatment.^17^ The subscores of FMA, Stroke Impairment Assessment Scale (SIAS)^18^, Action Research Arm Test (ARAT)^19^, Motor Activity Log-14 (MAL-14)^20^, Goal Attainment Scale (GAS)^21^, Barthel Index (BI)^22^, Modified Ashworth Scale (MAS)^23^, Stroke-Specific Quality of Life Scale (SS-QOL)^24^, were analyzed as secondary efficacy outcomes (Detailed information about each metric in appendix). In addition, the EEGs during exercise in both REAL and CTRL group were recorded and stored in the EEG-BCI based neurofeedback system. We compared changes in the success rate during the intervention between the REAL and CTRL groups using the difference between day 1 and day 10 data. Surface electromyogram (EMG) signals were measured during repetition of rest and extension. Modulation index (MI) at the first contraction and averaged MI across five repetitions were used as outcome measures (Appendix).

The safety primary endpoints in this study were proportions of adverse events and device-related problems. The number of events and the proportion will be calculated, and 95% confidence interval (CIs) for the proportions were estimated with the Clopper-Pearson method.

### Statistical analyses

To determine the sample size, average changes in FMA scores were assumed to be 6.9 and 3.0 for the REAL and CTRL group, respectively according to the reported previous reports. To reach a power of 80% with a type I error of 5%, 34 patients (17 patients per group) had to be included. Considering 15% exclusions from the analysis, the target sample size in this trial was set at 20 patients per group (total of 40 patients). The data were analyzed according to CONSORT statement.^25^ We assessed baseline descriptive statistics for all randomly assigned patients (the intention-to-treat population) by treatment allocation.

The hypothesis that BCI-based EEG neurofeedback is superior to standard therapy at 28 days post intervention was tested for the primary outcome (FMA score) at the two-sided significance level of 0·05. For this purpose, we performed a random effect ANCOVA for each endpoint, with group as fixed factors and the baseline FMA scores as a covariate. As a secondary analysis, we compared the prespecified secondary outcome allowing heteroscedasticity and autocorrelation. In addition, FMA scores were analyzed by another random effect ANCOVA model for each endpoint, with group as fixed factors and the baseline FMA scores, age, sex, type of stroke, lesion side, time since stroke, BCI accuracy on the first day of training as covariates. These results were examined exploratorily. The reasons for selecting the above covariates in the covariance model are described below. First, age and sex were selected as general demographic characteristics. For age, we rounded to the decade rather than per year to prevent overfitting the model. It was assumed that our non-stratified randomization could result in demographic differences and adding demographic variables as covariates would account for that. Second, we selected the type of stroke (infarction or hemorrhage), lesion side (right or left), lesion location (whether the damaged area overlaps with the CST), and time since stroke (3-6 months or >6 months) as the primary disease characteristics as has been reported in many BCI RCTs.^8,26,27^ Injury to the corona radiata or posterior limb of the internal capsule with overlapping damaged sites with CST is known to have a poor prognosis after stroke.^28,29^ Additionally, we categorized the period into 3 to 6 months and 6 months or longer since it is reported that spontaneous functional recovery is seen up to about 6 months after stroke onset ^30,31^ and many previous studies define the chronic phase as 6 months or longer. Lastly, FMA and BCI success rates on the first day of training were added as covariates for individuals’ baseline functions and abilities. The intention-to-treat population was included in both primary and secondary outcome analyses.

### Role of the funding source

The funding source did not have any involvement or influence in the study design.

## Results

Between March 30, 2017, and August 29, 2018, 40 eligible patients were enrolled by the participating practices, of whom 19 (47•5%) were allocated to REAL group and 21 (52·5%) to CTRL group, respectively (Figure 2). 19 (100%) patients in the REAL group and 21 (100%) patients in the CTRL group completed the treatment. The number of cases in the REAL and CTRL groups by facility was 10:10 in the Keio University Hospital (main hospital), 3:4 in Tokyo Bay Rehabilitation Hospital, 2:2 in Tokyo Metropolitan Rehabilitation Hospital, and 4:4 in Saiseikai Kanagawa Prefecture Hospital. Exercises were successfully conducted as planned, and no discontinuation cases were found in both groups. Patient demographic and clinical baseline characteristics are described in table 1. We found no between-group difference for parameters used to adjust the baseline (appendix).

**Table 1.**
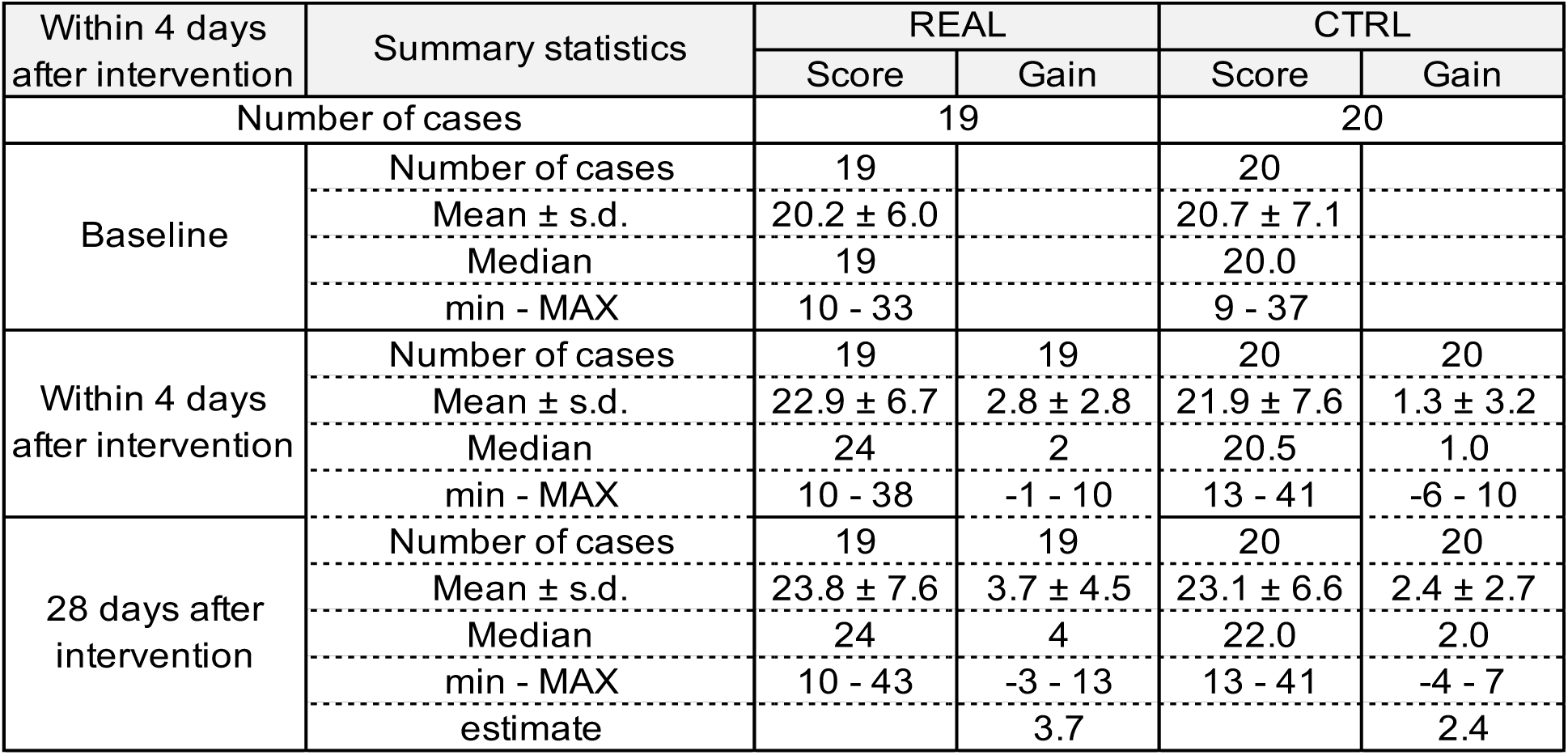
Summary of total FMA score.

| Within 4 days<br>after intervention | Summary statistics | REAL |  | CTRL |  |
| --- | --- | --- | --- | --- | --- |
|  |  | Score | Gain | Score | Gain |
| Number of cases |  | 19 |  | 20 |  |
| Baseline | Number of cases | 19 |  | 20 |  |
|  | Mean ± s.d. | 20.2 ± 6.0 |  | 20.7 ± 7.1 |  |
|  | Median | 19 |  | 20.0 |  |
|  | min - MAX | 10 - 33 |  | 9 - 37 |  |
| Within 4 days<br>after intervention | Number of cases | 19 | 19 | 20 | 20 |
|  | Mean ± s.d. | 22.9 ± 6.7 | 2.8 ± 2.8 | 21.9 ± 7.6 | 1.3 ± 3.2 |
|  | Median | 24 | 2 | 20.5 | 1.0 |
|  | min - MAX | 10 - 38 | -1 - 10 | 13 - 41 | -6 - 10 |
| 28 days after<br>intervention | Number of cases | 19 | 19 | 20 | 20 |
|  | Mean ± s.d. | 23.8 ± 7.6 | 3.7 ± 4.5 | 23.1 ± 6.6 | 2.4 ± 2.7 |
|  | Median | 24 | 4 | 22.0 | 2.0 |
|  | min - MAX | 10 - 43 | -3 - 13 | 13 - 41 | -4 - 7 |
|  | estimate |  | 3.7 |  | 2.4 |

For the ANCOVA model with the baseline FMA scores revealed that there is a significant main effect of time (*p* = 0·027) but not an interaction of time × intervention (*p* = 0·30) and main effect of intervention (*p* = 0·14). Post-hoc analysis revealed that significant differences were found between baseline and after-intervention period (*p* < 0·001), as well as baseline and follow-up period (*p* < 0·001). However, no between-group differences were found at any of time points.

In the secondary analysis with another ANCOVA model, we found a significant interaction of time × intervention for the primary outcome measure, that is FMA score (Table1, Figure 3, *p* = 0·037, η^2^ = 0·10). Post-hoc analysis revealed that REAL group exhibited a significant difference between the baseline and the follow-up period (*p* < 0·001) as well as the after-intervention period (*p* < 0·001). Moreover, there was statistically significant between-group differences at the after-intervention period (*p* = 0·027) while no differences were found at the baseline and follow-up period. The validity of the statistical model and effects of covariates on the treatment effect is explored in appendix.

**Figure 3.**
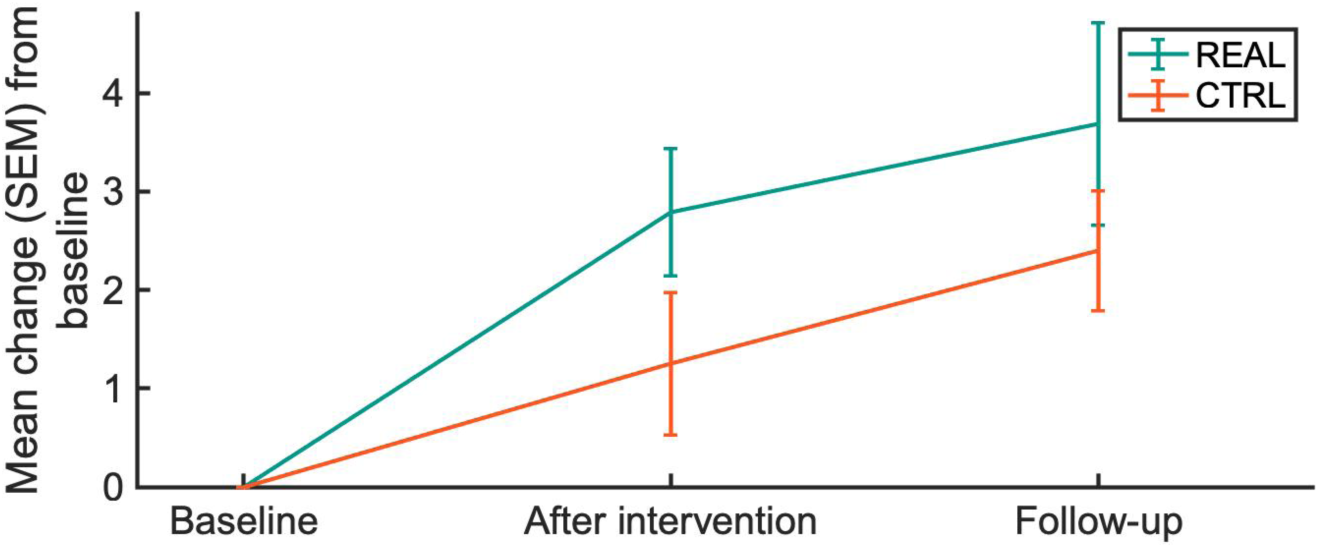
FMA time course in REAL and CTRL groups. The mean changes from baseline were visualized for each group.

Secondary outcome measures, such as, ARAT, MAL-14, GAS, BI, MAS, SS-QOL, were not statistically significant at any time points and between groups. For EEG signals, the REAL group significantly improved success rate of BCI control from day 1 to day10 (*p* = 0·036) relative to the CTRL group. For EMG signals, the REAL group significantly improved MI at the first trial and average MI across trials at follow-up period compared to baseline (*p*=0·018, *p* = 0·042, respectively).

Based on the Medical Dictionary for Regulatory Activities Terminology (MedDRA) by the International Council for Harmonisation of Technical Requirements for Pharmaceuticals for Human Use (ICH), adverse events reported during exercise were aggregated by System Organ Class (SOC) and Preffered Terms (PT). Adverse effects were 7 cases in the REAL group and 4 cases in the CTRL group. Dermatitis, irritation, and pain where the medical device contacts were reported for each case in the REAL group, and dermatitis where the sham device (same as the medical device, but robot-orthosis was not used and not activated) contacts was also reported in the CTRL group. In the REAL group, diarrhea, rashes, arthritis, and rashes were also reported. In the CTRL group, stomatitis, nasopharyngitis, and muscle pain were reported.

The number of adverse events that could not be ruled out as causally related to the investigational device was 3 in the REAL group (dermatitis, irritation, and pain) and 1 in the CTRL group (dermatitis). There were no adverse events that could not be ruled out as causally related to the interventional exercises. One adverse event, irritation at the contact of the medical device was found in the follow-up period of the CTRL group. All the events were mild, and there were no cases categorized into moderate or severe.

## Discussion

In the present trial, we show that motor imagery exercise combined with BCI and standard training can improve upper limb motor ability in patients with severer upper limb dysfunction after stroke. Although, we found no significant difference between patients allocated to REAL and CTRL groups in the first analysis, our exploratory analysis of the primary outcome measure considering patient heterogeneity showed that there is a significant interaction of time and intervention, indicating the FMA scores were significantly better in the REAL group than in the CTRL group. Moreover, we did not find any severe adverse effect throughout the trial, suggesting the clinical effectiveness of the BCI therapy.

To date, 12 RCTs on EEG-based BCI neurofeedback have been reported, all of which have shown efficacy. Although, no intervention for the chronic and severe segment has existed to date, four meta-analyses for the BCI neurofeedback have been reported, all with moderate-to-large effect sizes. ^8,26,27,32^ In this trial, we sought to confirm recovery of motor function of the paralyzed hand in this most treatment-resistant segment and compared with the effective treatment combination, that is the combination with motor imagery therapy and standard training.

First, for total FMA, scores increased with treatment. The recovery gain was marginal for MCID, but clinically adequate, since MCID in the chronic and severe segment can be considered lower than typical values. The superiority of MCID over the control group was observed in patients with a certain degree of preserved proximal function, suggesting that the treatment is more likely to be effective when there is some degree of functional cortico-muscular connectivity remaining.

Medical devices used in clinical trials were manufactured in accordance with Japanese domestic medical device standards (except for some items related to electrical safety that do not directly affect the trial, such as the strength of connectors and plastic cases). Although there was a total replacement of the robot-orthosis cable during the course of the study, the replacement was due to a durability problem with the connector, and did not affect the efficacy or safety of the clinical trial. No moderate and serious adverse events were reported in the clinical trial, and the trial was conducted as scheduled with no interruptions. Adverse events that could not be denied to be causally related to the medical device were dermatitis, irritation, and pain, but all were judged to be mild, and sufficient safety was confirmed.

Because of the design of this trial, in which one of the standard treatments, exercise imagery therapy, was used as a control group, the differences in intervention were discernible to the patients (PEDro scale in appendix). Also, for ethical considerations, patients in the CTRL group were informed in advance that they would receive BCI neurofeedback exercise after the follow-up period, and almost all patients chose this option. Patient motivation bias due to these factors cannot be denied.

Since the trial controlled for daily exercises of 40 minutes in duration, we were not able to dose control for the number of trials. In addition, we did not count any movement failures due to impedance changes or noise contamination during training, and the effects of these factors are unknown. Despite these issues, the superiority of EEG-based BCI neurofeedback over the control group in terms of treatment efficacy suggests that EEG-based BCI neurofeedback is a practical technique that can capture the neurological state and induce functional recovery.

## Conflicts of Interest

J.U. is a founder and representative director of the university startup company, LIFESCAPES Inc. involved in the research, development, and sales of rehabilitation devices, including brain-computer interfaces. He receives a salary from LIFESCAPES Inc. and holds shares in LIFESCAPES Inc. M.H. and R.Y are employed by LIFESCAPES Inc and receive a salary from LIFESCAPES Inc. S.I receives a salary from LIFESCAPES Inc. This company does not have any relationships with the device or setup used in the current study. The remaining author declare no competing interests.

## Data Availability

All data produced in the present study are available upon reasonable request to the authors

## Acknowledgements

The authors thank Shoko Tonomoto, Aya Kamiya and Sayoko Ishii for their assistance.

## Notes

### Clinical Trial

UMIN000026372

### Clinical Protocols

https://www.researchprotocols.org/2018/12/e12339/

### Funding Statement

This study was funded by AMED under Grant Number JP18hk0102032 and JSPS KAKENHI Grant Number 20H05923.

### Author Declarations

Ethics approval was granted by the Institutional Review Board of Keio University Hospital, Japan (D16-03, 2022-1092).

