## Appendix for "Hand motor recovery from post-stroke chronic severe hemiplegia with brain-computer interface based neurofeedback exercise: A randomized controlled trial"

**Supplementary methods**

*Apparatus*

All the components were functionally linked as follows (Figure S1) and medical device specification shown in Table S1.

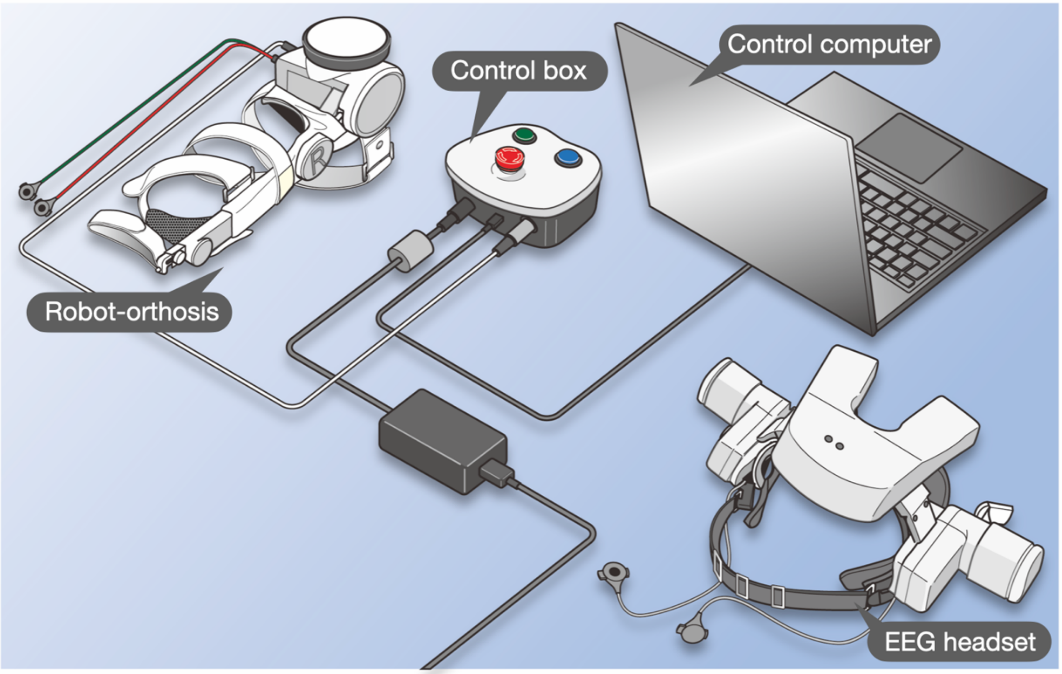

**Figure S1** EEG-based BCI neurofeedback apparatus. EEG headset records Scalp EEG over the ipsilesional sensorimotor cortex, and transmit to the control computer through wireless Bluetooth protocol in real-time. Control computer detects motor-related EEG response, known as the event-related desynchronization (ERD) of the sensorimotor rhythm (SMR), and sends control signals for neuromuscular electrical stimulation and robot-orthosis to the control box. The control box receives control signals from the control computer, and triggers actions of NMES and robot-orthosis. Also, the control box is used to operate finger extension/flexion, and start/stop of neurofeedback exercise, irrespective of EEG-based neurofeedback action. The control box generates electrical stimulation in response to the control signal from the control computer. The robot-orthosis is to be fastened tightly to patient’s paretic finger and forearm and extend/flex metacarpophalangeal joints of the fingers. The robot-orthosis equips the terminal of NMES.

**Table S1 Medical device specification**

| Items | | | Specification |
|---|---|---|---|
| EEG headset | Electrical ratings | Power supply | DC 2.4V (AAA 700 type; Nickel-Metal Hydride battery; 2 pieces) |
| | | Electrical protection | Internal Power Supply Equipment Type B Mounting Section |
| | Size | | W244 x H228 x D116 mm |
| | Weight | | 400 g |
| | Operating time | | 6 hours (when fully charged, 750 mAh) |
| | Electrode placement | | C3 (left hemisphere) and C4 (right hemisphere) in the International 10-20 methods and their 20 mm lateral |
| Control Compute | Electrical ratings | Power supply | DC 7.6 V (internal buttery supply) |
| | | Electrical protection | Internal Power Supply Equipment |
| Control Box | Electrical ratings | Power supply | DC 7.6 V (internal buttery supply) |
| | | Electrical protection | Internal Power Supply Equipment |
| | Size | | W148 x H75 x D145 mm |
| | Weight | | 1,000 g |
| Robot-orthosis | Electrical ratings | Power supply | AC 100 V (supplied from Control Box) |
| | | Electrical protection | Class 1 Equipment Type B Mounting Section |
| | Weight | | 500 g |
| | Cable length (connection to Control Box) | | 1.8 m |
| | Cable length (connection to NMES electr | | 0.15 m |

Prior to conducting the clinical trial, the following criteria (Tables S2 and S3) were verified to ensure that the equipment used met the medical device standards in Japan.

### Table S2 Validation of Validity

| Standards for validity | | Standards | Test procedure |
|---|---|---|---|
| Sensing | EEG measurement performance | Compliant with IEC 6061-2-26:2012 201.12.1.101.1 (Accuracy of signal reproduction) | Same as the definition on the left. |
| | | Compliant with IEC 6061-2-26:2012 201.12.1.101.2 (Input dynamic range and differential offset voltage) | Same as the definition on the left. |
| | | Compliant with IEC 6061-2-26:2012 201.12.1.101.3 (Input noise) | Same as the definition on the left. |
| | | Compliant with IEC 6061-2-26:2012 201.12.1.101.4 (Frequency response) | Same as the definition on the left. |
| | | Compliant with IEC 6061-2-26:2012 201.12.1.101.5 (Common mode rejection) | In the definition on the left, the input signal "A 1 Vr.m.s. signal at mains frequency" was changed to"A 1 Vpp signal at mains frequency" and inspection was conducted. |
| Detection | EEG-SMR-ERD detection performance | Calculated ERD level must be within 19±5% in any 4 signals. | Connects 30 k ohm registance between EEG headset electrode section and functional generator, and output 4 types of signals that satisfy ERD level being 19% from the functional generator to EEG headset. In each type of test signals, the 3-second average of the ERD level measured by EEG headset and Control Computer is used. |
| Actuation | Robot-orthosis performance | The device must be able to flex patient's metacarpophalangeal joint up to 80 degrees to the palmar flexion side, with the extension position of the joint at 0 degrees. | Measure the angle between the top surface of the palm side pipe receiver and the top surface of the rotating arm at the maximally extended position with a protractor. |
| | Neuromuscular electrical stimulation performanc | The electrical pulse must be bipolar recutangular waveform of which pulse duration is 1 ms. | Connect 1 k ohm registance between the anode and cathode connector of the NMES cable, and observe output signal through an oscilloscope. The cycle of the observed volutage signal must be 10 ms, and the polarity should be positive in 0-1 ms and negative in 1-2 ms, followed by 0 V in 2-10 ms. |

#### Table S3 Validation of safety

| Standards for safety | Standards | Test procedure |
|---|---|---|
| Basic safety<br>Electrical safety<br>Mechanical safety | (Dec 1 2016-May 12 2017) Compliant with JIS T 0601-1:1999 Medical Electrical Equipment Part 1: General requirements for satefy. | Same as the definition on the left. |
| | (from May 13 2017) Compliant with JIS T 0601-1:2012 Medical Electrical Equipment Part 1: General requirements for basic satefy and general performance. | Same as the definition on the left. |
| Specific requirements for basic safety and basic performance of nerve and muscle stimulators | Compliant with JIS 0601-2-10:2015 Medical Electrical Equipment Part 2-10: Specific requirements for basic safety and basic performance of nerve and muscle stimulators. | Same as the definition on the left. |
| Electromagnetic compatibility (EMC) | Compliant with JIS T 0601-2-1-2:2012 General requirements for satefy, requirement for electromagnetic compatibility and testing. | Same as the definition on the left. |
| Programmable electrical medical system (PEMS) | Compliant with JIS T 0601-1:2012 Medical Electrical Equipment Part 1: General requirements for basic satefy and general performance. | Same as the definition on the left. |
| Biological safety | Compliant with JIS 0993-1:2012 Biological evaluation of medical devices Part 1: Evaluation and testing in the risk management process. | |

| Standards for safety (continued) | Standards | Test procedure |
|---|---|---|
| Safety for robot-orthosis (Safety devices and alarm functions) | The movable range of the finger-loading part shall be limited from 0 to 80 degrees by the mechanical stopper. The mechanical stopper must not break even under a load of 11 kg (twice the maximum load). | Confirm in the drawing that the movable range of the finger-loading part is limited from 0 to 80 degrees by the mechanical stopper. Confirm that the mechanical stopper is not damaged by a load of 11 kg or more. |
| | After detecting that the finger-loading part has not reached the set target angle on the extended side during the extension operation, the finger-loading part shall return 25 degrees within 5 seconds. | Set the movement speed of the motor part to 10.5 degrees per second, set the target angle for the extension side of the finger-loading part to 80 degrees, and extend the finger-loading part from the flexion side to the extension side. During the extension movement, mechanically lock the finger-loading part at an angle of 60 degrees, and confirm that the angle of the finger rest returns to the flexion side 25 degrees from the target angle set for the extension side within 5 seconds after locking (the angle of the finger-loading part is 75 degrees). |
| | The system shall be equipped with an emergency stop button and shall be able to be shut down by the emergency stop button. | During the movement of the robot-orthosis, press the emergency stop switch and confirm that the voltage and electrical stimulation output of the motor is less than 0.2C. |
| | If the motor continues to run for 300 seconds, the motor operation shall be stopped. | Confirm that the operating voltage of the motor is 0.2V or less after 300 seconds ±10% of continuous motor operation. |

*Electroencephalogram Recording*

Ag-AgCl electrodes (φ=9 mm) for EEG measurement was placed over the ipsi-lesional sensorimotor cortex, namely, C3 (for the left hemisphere) or C4 (for the right hemisphere) according the international 10-20 system. An additional electrode was placed 20 mm lateral to C3 or C4. A ground electrode was placed on A1, and the reference electrode was placed on A2. All electrodes were guided manually and fixed with a custom-made EEG headset. The application-specific integrated circuit–based analog circuit and microprocessor were embedded inside the headset, and 2-channel EEGs were derived in a monopolar manner and processed with ×1,200 amplification and 0.21 to 199 Hz filtering. The processed EEG signals were digitized at 200 Hz with 12 bits (least significant bit 0.366 μV). Note that a notch filter of 50 Hz was used to minimize the power-line noise. EEGs were then transmitted to a laptop using a Bluetooth 3.0 wireless protocol and were subtracted from each other to derive a bipolar EEG. A 2 Hz to 50 Hz bandpass filter with a 50 Hz notch was again used to reduce noise contamination.

A 1-second time-sliding Hanning window was applied to this bipolar EEG signal with 87.5% overlap, and fast Fourier transform was applied to obtain the time-varying power spectrum of the signal. The two-dimensional (2D) feature vector with mean alpha frequency band power (7-13 Hz) and mean beta frequency band power (14-26 Hz) was constructed at each time segment and traced with time in the feature space. The discriminant line that determines EEG feature vectors as in either the *ERD* or *baseline* class was used for EEG labeling. The discriminant line was calibrated for each participant every day before the training session (see also Calibration section below).

*Estimation of Sensorimotor Cortex Excitability from Electroencephalogram*

Alpha and its harmonic beta frequency band powers in EEG recorded over the sensorimotor cortex is called as the Sensorimotor Rhythm (SMR). Rodents, monkeys, and human studies have evident its neurobiological substrate of the inhibitory recurrent relay cells in the thalamo-cortical loop. A coupled and synchronized cortical population activity is formed by the pulling activity of this circuit during the resting state, but is disrupted during motor imagery or execution if the thalamo-cortical circuits desynchronously elevate their excitability. Event-Related Desynchronization (ERD), the amplitude attenuation, of the EEG-SMR is therefore often observed during motor imagery or execution. EEG-SMR-ERD is now a known analog of the increased sensorimotor cortex excitability, associated with disinhibition of gamma-aminobutyric acid-ergic intracortical inhibitory circuits [8]. It is also correlated with the increase of the corticospinal tract excitability and the spinal anterior horn cell excitability [9] with an agonist muscle specific. Extrapolation of these findings to poststroke patients with hemiplegia may be acceptable because EEG-SMR-ERD during paretic hand motor imagery is associated with ipsi-lesional corticospinal tract excitability in poststroke patients with hemiplegia [38].

Alpha frequency oscillation and its resonance among cortical and subcortical regions in poststroke patients with hemiplegia predicted motor outcome, suggesting that the alpha component is related to sensorimotor function. Recent clinical studies with EEG-based BCI neurofeedback exercise intervention also suggest that up-conditioning of EEG alpha and beta band frequency powers and their ERD during motor attempting of paretic finger opening through EEG-based BCI neurofeedback exercise is associated with increased corticospinal tract excitability [10] and the blood oxygen level-dependent signal of magnetic resonance imaging in the ipsilesional sensorimotor cortex [11,12]. Repeated use of EEG-BCI based neurofeedback during mental practice of motor imagery that forces patients to increase alpha and beta frequency powers at rest and decrease their power during paretic hand motor attempting is, therefore, ensure a high compliance level of the voluntary recruitment of the remaining

*Calibration procedure of the EEG-BCI based neurofeedback*

A 2D feature vector with mean alpha frequency band power (7-13 Hz) and mean beta frequency band power (14-26 Hz) was first obtained from a 1-second time-sliding Hanning window with 87.5% overlap applied to the collected EEG data.

The feature vectors with annotations of either "imagine paretic finger extension" or "keep relaxing" were mapped onto the feature space, and the parameters in the LDA algorithm were optimized to separate the features into appropriate classes. Consequently, the LDA in the EEG-BCI based neurofeedback exercise becomes to return a value of +1 (the sensorimotor cortex excitability is the resting level) or −1 (the sensorimotor cortex excitability is increased from the resting level) every 125 ms according to the EEGs. Noted here that neither NMES nor robot-orthosis action were given during calibration session, irrespective to EEG response.

*Procedure of assessment*

The primary and secondary outcomes were acquired in the following schedule (Table. S4). The blind evaluator acquired the primary outcome.

**Table S4** Schedule of assessment

| Time points and measure | Baseline | Intervention ( 10 session in 2 weeks) | Posttreatment (within 4 days after treatment) | Follow-up (28 days after treatment) |
|---|---|---|---|---|
| **Primary outcome measure** | | | | |
| Fugl-Meyer Assessment | B | – | B | B |
| **Secondary outcome measure** | | | | |
| Action Research Arm Test | E | – | E | E |
| Motor Activity Log-14 | E | – | E | E |
| Stroke Impairment Assessment Set | E | – | E | E |
| Modified Ashworth Scale | E | – | E | E |
| Barthel Index | E | – | – | E |
| Goal Attainment Scale | S | – | – | S |
| Stroke-Specific Quality of Life Scale | S | – | – | S |
| Surface Electromyogram | EP | | EP | EP |
| Electroencephalogram during excercises | – | EP | – | – |

B: Assessment of blided evaluator. -： Not applicable. E: Assessment of well-trained evaluator. S: Participant self-report. EP: Electrophysiological data.

The secondary outcome measures were prospectively determined to evaluate the effect of intervention in a variety of aspects. Each metric represents the following sensorimotor abilities:

*Action Research Arm Test*

Action Research Arm Test (ARAT) [40] is a frequently used, validated, and reliable measure of upper extremity function with 4 subsections: grip, grasp, pinch, and gross movement [41,42]. The maximum summed score is 57.

*Motor Activity Log-14*

Upper extremity disability in activities of daily living (ADL) was assessed with Motor Activity Log (MAL), which uses a structured interview [43]. MAL includes 14 items, scored on an 11-point amount of use scale (range 0-5) to rate how much the arm is used (MAL-amount of use) and an 11-point quality of movement scale (range 0-5) to rate how well the participants are using their affected upper extremity [43]. High construct validity and reliability have been reported in patients with chronic stroke [43,44].

*Motor Scores of the Stroke Impairment Assessment Set*

The Stroke Impairment Assessment Set (SIAS) is a comprehensive instrument for assessing stroke impairment with well-established psychometric properties [45,46]. SIAS assesses various aspects of impairment in stroke patients, including motor function, tone, sensory function, range of motion, pain, trunk function, visuospatial function, speech, and sound side function. Motor scores of the SIAS are composed of 5 items that assess arm, finger, hip, knee, and ankle functions and are rated from 0 (severely impaired) to 5 (normal).

*Goal Attainment Scale*

The Goal Attainment Scale (GAS) is a self-rating scale to evaluate subjective improvement following rehabilitation [47-50]. Patients rate the attainment level of the rehabilitation outcome for the goal that they set themselves. If the attainment is as expected, it is rated as 0. Improvement beyond expectation is rated +1 or +2 and that below expectation is rated −1 or −2.

*Barthel Index*

The Barthel Index (BI) is one of the most frequently used measures to evaluate ADL in stroke research [51,52]. BI measures independence in ADL; the maximum score is 100. The 10 assessed items of ADL are feeding, bathing, grooming, dressing, bowel control, bladder control, toilet use, transfers, mobility, and ascending and descending stairs.

*Modified Ashworth Scale*

Spasticity at the wrist and finger flexors of the affected upper extremity was assessed with the Modified Ashworth Scale (MAS), a 6-point rating scale used to measure passive muscle resistance [53].

*Stroke-Specific Quality of Life Scale*

The Stroke-Specific Quality of Life Scale (SS-QOL) was developed to assess health-related quality of life in stroke patients [54,55]. SS-QOL contains 49 items and covers 12 different areas of quality of life affected by stroke. The 12 areas of the SS-QOL are energy, family roles, language, mobility, mood, personality, self-care, social roles, thinking, upper extremity function, vision, and work or productivity. Each area can be scored separately, but a total score is also available. The possible range of all scales is from 1 to 5, where a lower value indicates a lower health-related quality of life.

*Surface Electromyogram*

The muscle activities of the paretic extensor digitorum communis and flexor digitorum superficialis muscles were recorded with Ag-AgCl surface electrodes with diameters of 9 mm (Nihon Kohden, Tokyo, Japan). The electrodes were applied with center-to-center spacing of 30 mm and were placed parallel to the muscle fibers and distal from the motor points of individual muscles. Before the electrodes were attached, the skin areas were rubbed with alcohol. Skin resistance was kept below 5 kΩ. An MEB-2300 EMG machine (Nihon Kohden) was used to record and analyze the EMG data. The bandpass filter was set at 20 Hz to 1 kHz. The patients were seated in a comfortable chair with their arms on an armrest and the angle of their elbows was kept at 70 to 90 degrees. They were instructed to rest for 5 seconds and then to extend their affected fingers for the next 5 seconds for 1 cycle. In total, 5 cycles of 5 seconds of rest and 5 seconds of extension were repeated.

*Electroencephalogram during exercise*

The EEGs during exercise in both REAL and CNTRL group were recorded and stored in the EEG-BCI based neurofeedback system. We compared the magnitude and duration of ERD between the REAL and CTRL groups.

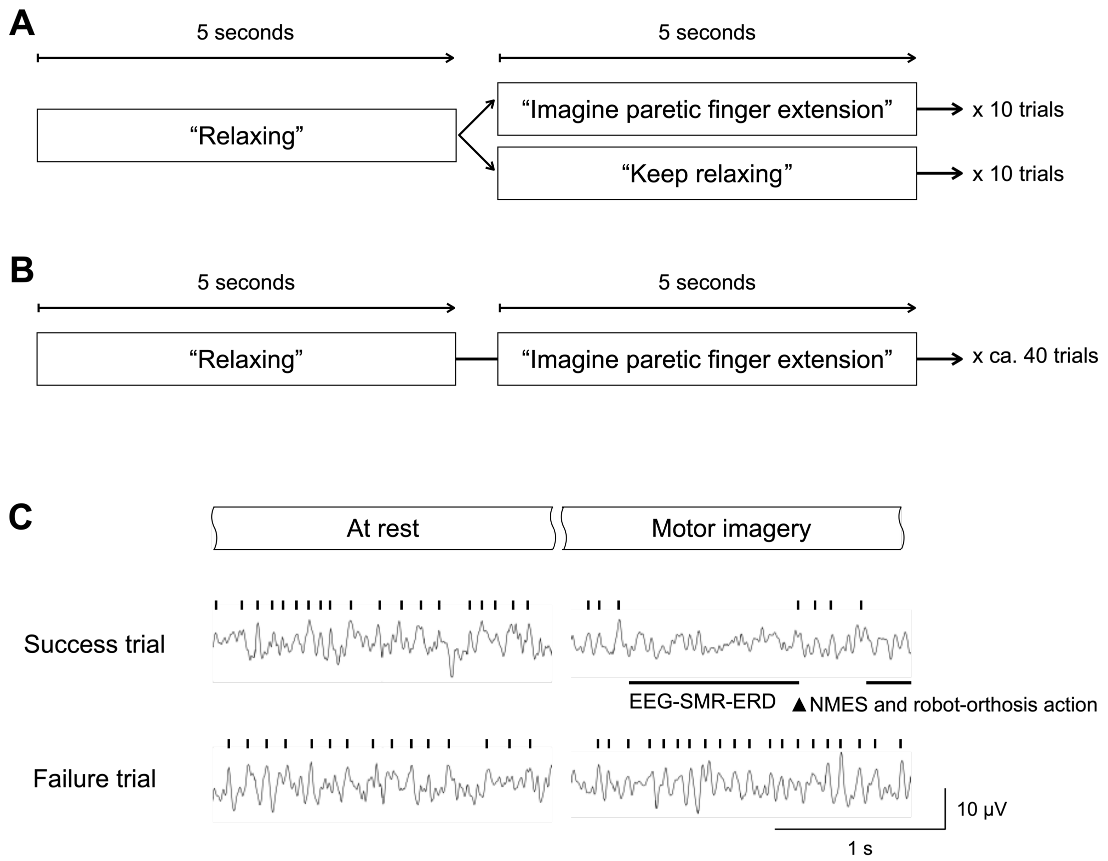

**Figure S2** Time-course of neurofeedback. (A) Time course in the calibration session. Either motor imagery or keeping rest is randomly cued. (B) Time course in the exercise session. After a 5-sec rest period, patients were asked to perform kinesthetic motor imagery of finger extension at the affected side in 5 seconds. The entire exercise time was set 40 min., and therefore the exercise consisted of ca 40 trials per day. (C) Time course of system action. If EEG-SMR (shown as vertical bar) seen in the resting-state is disappeared and EEG-SMR-ERD was observed during motor imagery period for more than 1 sec., the system triggers NMES and robot-orthosis action (upper trace). If EEG-SMR-ERD was not seen, no system action was happened (lower trace).

*Outcome Measurement Schedule of Assessment*

The primary outcome measure was assessed by a blinded evaluator. Other functional measurements were assessed by evaluators who were trained by the organizer of this RCT. Most assessments were conducted at baseline, after intervention (post), and 4 weeks after intervention (follow-up). EEGs were recorded by the EEG-BMI rehabilitation system during each training session. The schedule of assessments is shown in Table S5.

**Table S5** Schedule of assessment

| Time points and measure | Baseline | Intervention ( 10 session in 2 weeks) | Posttreatment (within 4 days after treatment) | Follow-up (28 days after treatment) |
|---|---|---|---|---|
| **Primary outcome measure** | | | | |
| Fugl-Meyer Assessment | B | – | B | B |
| **Secondary outcome measure** | | | | |
| Action Research Test | E | – | E | E |
| Motor Activity Log-14 | E | – | E | E |
| Stroke Impairment Assessment Set | E | – | E | E |
| Modified Ashworth Scale | E | – | E | E |
| Barthel Index | E | – | – | E |
| Goal Attainment Scale | S | – | – | S |
| Stroke-Specific Quality of Life Scale | S | – | – | S |
| Surface Electromyogram | EP | | EP | EP |
| Electroencephalogram during excercis | – | EP | – | – |

B: Assessment of blided evaluator. -：Not applicable. E: Assessment of well-trained evaluator. S: Participant self-report. EP: Electrophysiological data.

*EMG analysis*

To assess whether the BCI intervention improved voluntary contraction and relaxation switching and its repetition from surface EMG data, the Modulation index was calculated as follows: (1) acquired raw EMG signals over EDC and FDS muscles underwent a 20–500 Hz second-order Butterworth bandpass filter and 49–51 Hz, and 99–101 Hz second-order notch filters; (2) filtered EMG data was segmented into 5 trials, with 1 trial was consisted of 5 s of the resting epoch and 5 s of the task (finger extension) epoch; (3) the RMS values were calculated (100 ms segments with 90% overlap) by cutting out 1 s in the middle of the resting and task epochs, respectively; (4) the median of the RMS values in the resting and task epochs were obtained; and (5) the Modulation index was calculated from the following formula:

$$MI(t) = \frac{A(t)}{R(t)}$$

where MI is the modulation index over EDC or FDS muscles, A(t) is the median RMS value in the task epoch, and R(t) is the median RMS value in the resting epoch. The switching ability from relaxation into voluntary contraction was evaluated from the MI value in the first trial, and its repetition capability was examined in the average of total a of five trials.

In addition, the co-contraction index (CCI) was calculated from the EDC and FDS EMGs to determine whether the agonist muscles became selectively activated during finger extension. The changes in the Modulation index CCI from baseline (i.e., DAY1) on the last day of BCI-neurofeedback treatment were compared between treatment groups by Wilcoxon's rank sum test, respectively.

##### *Physiotherapy Evidence Database scale*

The Physiotherapy Evidence Database (PEDro) scale to assess the internal validity of the present study was shown in Table S6. Eight out of eleven sections were labeled as “Yes”.

**Table S6** Physiotherapy Evidence Database (PEDro) scale

| | | |
|---|---|---|
| 1 | Eligibility Criteria | Yes |
| 2 | Random Allocation | Yes |
| 3 | Concealed Allocation | Yes |
| 4 | Baseline Comparability | Yes |
| 5 | Blind Assessors | Yes |
| 6 | Blind Subjects | No |
| 7 | Blind Therapists | No |
| 8 | Adequate Follow Up | Yes |
| 9 | Intention-to-Treat Analysis | Yes |
| 10 | Between-Group Comparisons | Yes |
| 11 | Point Estimates and Variability | Yes |

#### Supplementary Results

*Patient Characteristics*

Patient characteristics were assessed to ensure the between-group homogeneity (Table S7). Summary statistics were calculated for each demographic variables (sex, age, weight, height) and disease-related variables (primary disease, complications, concomitant medications). There were no notable differences that could affect effectiveness and safety in both groups.

**Table S7** Demographic of patients by treatment group

| Characteristics items | | FAS | | PPS | | Population for safety analysis | |
|---|---|---|---|---|---|---|---|
| | | REAL | CTRL | REAL | CTRL | REAL | CTRL |
| Subject to analysis | | 19 | 20 | 19 | 20 | 19 | 21 |
| Sex | Male | 16 (84%) | 11 (55%) | 16 (84%) | 11 (55%) | 16 (84%) | 12 (57%) |
| | Female | 3 (16%) | 9 (45%) | 3 (16%) | 9 (45%) | 3 (16%) | 9 (43%) |
| Age | Mean ± s.d. | 54 ± 10 | 48 ± 13 | 54 ±10 | 48 ± 13 | 54 ±10 | 48 ±13 |
| | Median | 52 | 49 | 52 | 49 | 52 | 49 |
| | min - MAX | 35 - 76 | 18 - 65 | 35 - 76 | 18 -65 | 35 - 76 | 18 -65 |
| Weight | Mean ± s.d. | 66.0 ± 13.3 | 61.9 ± 15.4 | 66.0 ± 13.3 | 61.9 ± 15.4 | 66.0 ± 13.3 | 62.2 ± 15.1 |
| | Median | 65.4 | 60.0 | 65.4 | 60.0 | 65.4 | 61.8 |
| | min - MAX | 46.5 - 96.0 | 45.0 - 100.5 | 46.5 - 96.0 | 45.0 - 100.5 | 46.5 - 96.0 | 45.0 - 100.5 |
| Height | Mean ± s.d. | 168.8 ± 5.6 | 162.0 ± 10.9 | 168.8 ± 5.6 | 162.0 ± 10.9 | 168.8 ± 5.6 | 162.4 ± 10.8 |
| | Median | 169.7 | 163.8 | 169.7 | 163.8 | 169.7 | 164.5 |
| | min - MAX | 158.5 - 183.0 | 141.5 - 180.0 | 158.5 - 183.0 | 141.5 - 180.0 | 158.5 - 183.0 | 141.5 - 180.0 |
| Primary disease | Infarct | 7 (37%) | 5 (25%) | 7 (37%) | 5 (25%) | 7 (37%) | 5 (24%) |
| | Hemorrhage | 12 (63%) | 15 (75%) | 12 (63%) | 15 (75%) | 12 (63%) | 16 (76%) |
| | Subarchnoid hemorrhage | 0 (0%) | 0 (0%) | 0 (0%) | 0 (0%) | 0 (0%) | 0 (0%) |
| Complications | yes | 1 (5%) | 2 (10%) | 1 (5%) | 2 (10%) | 1 (5%) | 2 (10%) |
| | no | 18 (95%) | 18 (90%) | 18 (95%) | 18 (90%) | 18 (95%) | 19 (91%) |
| Comcomitant medications | yes | 0 (0%) | 1 (5%) | 0 (0%) | 1 (5%) | 0 (0%) | 1 (5%) |
| | no | 19 (100%) | 19 (95%) | 19 (100%) | 19 (95%) | 19 (100%) | 20 (95%) |

Motor function score at the baseline period before intervention was assessed. There were no notable differences in total FMA score and its subscores (A: shoulder-elbow-forearm, B: wrist, C: finger, and D: coordination and speed) that could affect effectiveness and safety in both groups (see Table S6). Also, there were no notable differences in SIAS subscores (Upper extremity (proximal), Fingers, Lower extremity (hip joint), Lower extremity (knee joint), Lower extremity (ankle joint)) that could affect effectiveness and safety in both groups (see Table S7). Individual patient demographic and baseline clinical characteristics are listed in Table S8.

**Table S8** Individual patient demographic and clinical characteristics

| Patients | Group | Days from stroke onset to baseline assesment | Sex | Age | Primary disease | | |
|---|---|---|---|---|---|---|---|
| | | | | | Type of stroke | Lesion | Paralyzied limb |
| 1 | REAL | 2,265 | Male | 40-49 | Haemorrhage | Right putamen | Left |
| 2 | CTRL | 1,548 | Female | 60-69 | Haemorrhage | Left putamen | Right |
| 3 | REAL | 223 | Male | 50-60 | Haemorrhage | N/A | Left |
| 4 | REAL | 978 | Male | 60-69 | Infarction | N/A | Left |
| 5 | CTRL | 291 | Female | 20-29 | Haemorrhage | Left frontal lobe | Right |
| 6 | REAL | 1,334 | Male | 50-59 | Haemorrhage | Left putamen | Right |
| 7 | CTRL | 1,204 | Female | 60-69 | Infarction | Right corona radiata | Left |
| 8 | REAL | 252 | Female | 60-69 | Infarction | Left corona radiata | Right |
| 9 | CTRL | 571 | Male | 40-49 | Haemorrhage | Left putamen | Right |
| 10 | CTRL | 511 | Male | 50-59 | Haemorrhage | Right thalamus | Left |
| 11 | CTRL | 3,332 | Female | 60-69 | Infarction | Right putamen～corona radiata | Left |
| 12 | REAL | 1,729 | Female | 50-59 | Infarction | Left anterior cerebral artery | Right |
| 13 | REAL | 356 | Male | 50-59 | Haemorrhage | Left putamen | Right |
| 14 | CTRL | 243 | Male | 40-49 | Infarction | Left putamen～corona radiata | Right |
| 15 | CTRL | 1,751 | Female | 60-69 | Haemorrhage | Right thalamus | Left |
| 16 | CTRL | 404 | Male | 40-49 | Infarction | Left corona radiata | Right |
| 17 | REAL | 390 | Male | 40-49 | Haemorrhage | Left putamen | Right |
| 18 | CTRL | 1,577 | Male | 40-49 | Haemorrhage | Left putamen | Left |
| 19 | REAL | 697 | Male | 50-59 | Haemorrhage | Left putamen | Right |
| 20 | CTRL | 3,803 | Female | 60-69 | Haemorrhage | Right basal ganglia | Left |
| 21 | CTRL | 885 | Male | 40-49 | Haemorrhage | Left thalamus | Right |
| 22 | CTRL | 96 | Male | 40-49 | Infarction | Left corona radiata | Right |
| 23 | REAL | 435 | Male | 30-39 | Haemorrhage | Right putamen | Left |
| 24 | REAL | 393 | Male | 40-49 | Infarction | N/A | Left |
| 25 | CTRL | 1,520 | Female | 50-59 | Haemorrhage | Right thalamus | Left |
| 26 | REAL | 135 | Male | 70-79 | Infarction | Left CA | Right |
| 27 | REAL | 4,879 | Male | 60-69 | Infarction | N/A | Right |
| 28 | CTRL | 812 | Male | 50-59 | Haemorrhage | Right putamen | Left |
| 29 | CTRL | 3,415 | Male | 40-49 | Haemorrhage | Left putamen | Right |
| 30 | REAL | 311 | Male | 30-39 | Haemorrhage | Left cortex | Right |
| 31 | REAL | 1,404 | Male | 70-79 | Infarction | Right middle cerebral artery | Left |
| 32 | CTRL | 420 | Male | 40-49 | Haemorrhage | N/A | Right |
| 33 | REAL | 1,557 | Male | 50-59 | Haemorrhage | Right putamen | Left |
| 34 | CTRL | 263 | Female | 40-49 | Haemorrhage | Left putamen | Right |
| 35 | REAL | 2,710 | Female | 50-59 | Haemorrhage | Left putamen | Right |
| 36 | CTRL | 334 | Female | 10-19 | Haemorrhage | Right subcortical frontal lobe | Left |
| 37 | CTRL | 3,831 | Male | 50-59 | Haemorrhage | Left putamen | Right |
| 38 | REAL | 904 | Male | 60-69 | Haemorrhage | Left putamen・Left subcortical frontal lobe | Right |
| 39 | REAL | 2,557 | Male | 50-59 | Haemorrhage | Left putamen | Right |
| 40 | CTRL | 128 | Male | 40-49 | Haemorrhage | Right putamen | Left |

*Primary Outcome*

The total and subscores of FMA were summarized in Table S9. The FMA scores suggested that scores at each time point in each group indicate the normality according to Shapiro-Wilk test (all $p$ > 0.05). The sphericity of variance was confirmed using Mauchly's test of sphericity (p = 0.108). The independence of covariates and independent variables are confirmed by the absence of statistically significant interaction between intervention and covariates ($p$ > 0.05).

ANOVA model used in the secondary analysis revealed that the intervention effect specific to the REAL was found in the patients with corticospinal tract (CST) injury and baseline FMA scores are significantly correlated with the treatment effects. The statistically significant regression models were not found after the family-wise error rate correction.

**Table S9** FMA score at the baseline period before intervention

| Characteristics items | | FAS | | PPS | | Population for safety analysis | |
|---|---|---|---|---|---|---|---|
| | | REAL | CTRL | REAL | CTRL | REAL | CTRL |
| Subject to analysis | | 19 | 20 | 19 | 20 | 19 | 21 |
| Total | Mean ± s.d. | 20.2 ± 6.0 | 20.7±7.1 | 20.2±6.0 | 20.7±7.1 | 20.2±6.0 | 20.2±7.2 |
| | Median | 19 | 20 | 19 | 20 | 19 | 20 |
| | min - MAX | 10 - 33 | 9 - 37 | 10 - 33 | 9 - 37 | 10 - 33 | 9 - 37 |
| A (Shoulder-elbow-foream) | Mean ± s.d. | 15.3 ± 5.8 | 16.8 ± 5.7 | 15.3 ± 5.8 | 16.8 ± 5.7 | 15.3 ± 5.8 | 16.5 ± 5.7 |
| | Median | 15 | 17.5 | 15 | 17.5 | 15 | 17 |
| | min - MAX | 8 - 26 | 8 - 28 | 8 - 26 | 8 - 28 | 8 - 26 | 8 - 28 |
| B (Wrist) | Mean ± s.d. | 0.7 ±1.6 | 0.7 ±1.5 | 0.7 ±1.6 | 0.7 ±1.5 | 0.7 ±1.6 | 0.7 ±1.5 |
| | Median | 0.0 | 0.0 | 0.0 | 0.0 | 0.0 | 0.0 |
| | min - MAX | 0 - 5 | 0 - 4 | 0 -5 | 0 - 4 | 0 -5 | 0 - 4 |
| C (Fingers) | Mean ± s.d. | 4.1 ± 1.9 | 3.2 ± 1.9 | 4.1 ± 1.9 | 3.2 ± 1.9 | 4.1 ± 1.9 | 3.0 ± 1.9 |
| | Median | 4.0 | 3.0 | 4.0 | 3.0 | 4.0 | 3.0 |
| | min - MAX | 1 - 7 | 1 - 6 | 1 - 7 | 1 - 6 | 1 - 7 | 1 - 6 |
| D (Cordination and speed) | Mean ± s.d. | 0.0 ± 0.0 | 0.0 ± 0.0 | 0.0 ± 0.0 | 0.0 ± 0.0 | 0.0 ± 0.0 | 0.0 ± 0.0 |
| | Median | 0.0 | 0.0 | 0.0 | 0.0 | 0.0 | 0.0 |
| | min - MAX | 0 - 0 | 0 - 0 | 0 - 0 | 0 - 0 | 0 - 0 | 0 - 0 |

*Secondary Outcome*

**Table S10** SIAS score at the baseline period before intervention

| Characteristics items | | FAS | | PPS | | Population for safety analysis | |
|---|---|---|---|---|---|---|---|
| | | REAL | CNTL | REAL | CNTL | REAL | CNTL |
| Subject to analysis | | 19 | 20 | 19 | 20 | 19 | 21 |
| Upper extremity (proximal) | 5 | 0 (0%) | 0 (0%) | 0 (0%) | 0 (0%) | 0 (0%) | 0 (0%) |
| | 4 | 0 (0%) | 1 (5%) | 0 (0%) | 1 (5%) | 0 (0%) | 1 (5%) |
| | 3 | 11 (58%) | 12 (60%) | 11 (58%) | 12 (60%) | 11 (58%) | 12 (57%) |
| | 2 | 8 (42%) | 7 (35%) | 8 (42%) | 7 (35%) | 8 (42%) | 8 (38%) |
| | 1 | 0 (0%) | 0 (0%) | 0 (0%) | 0 (0%) | 0 (0%) | 0 (0%) |
| | 0 | 0 (0%) | 0 (0%) | 0 (0%) | 0 (0%) | 0 (0%) | 0 (0%) |
| | Not evaluated/unknown | 0 (0%) | 0 (0%) | 0 (0%) | 0 (0%) | 0 (0%) | 0 (0%) |
| Fingers | 5 | 0 (0%) | 0 (0%) | 0 (0%) | 0 (0%) | 0 (0%) | 0 (0%) |
| | 4 | 0 (0%) | 0 (0%) | 0 (0%) | 0 (0%) | 0 (0%) | 0 (0%) |
| | 3 | 0 (0%) | 0 (0%) | 0 (0%) | 0 (0%) | 0 (0%) | 0 (0%) |
| | 2 | 0 (0%) | 0 (0%) | 0 (0%) | 0 (0%) | 0 (0%) | 0 (0%) |
| | 1c | 0 (0%) | 0 (0%) | 0 (0%) | 0 (0%) | 0 (0%) | 0 (0%) |
| | 1b | 0 (0%) | 0 (0%) | 0 (0%) | 0 (0%) | 0 (0%) | 0 (0%) |
| | 1a | 19 (100%) | 20 (100%) | 19 (100%) | 20 (100%) | 19 (100%) | 21 (100%) |
| | 0 | 0 (0%) | 0 (0%) | 0 (0%) | 0 (0%) | 0 (0%) | 0 (0%) |
| | Not evaluated/unknown | 0 (0%) | 0 (0%) | 0 (0%) | 0 (0%) | 0 (0%) | 0 (0%) |
| Lower extremity (hip joint) | 5 | 1 (5%) | 1 (5%) | 1 (5%) | 1 (5%) | 1 (5%) | 1 (5%) |
| | 4 | 5 (26%) | 8 (40%) | 5 (26%) | 8 (40%) | 5 (26%) | 8 (38%) |
| | 3 | 12 (63%) | 10 (50%) | 12 (63%) | 10 (50%) | 12 (63%) | 10 (48%) |
| | 2 | 1 (5%) | 1 (5%) | 1 (5%) | 1 (5%) | 1 (5%) | 2 (10%) |
| | 1 | 0 (0%) | 0 (0%) | 0 (0%) | 0 (0%) | 0 (0%) | 0 (0%) |
| | 0 | 0 (0%) | 0 (0%) | 0 (0%) | 0 (0%) | 0 (0%) | 0 (0%) |
| | Not evaluated/unknown | 0 (0%) | 0 (0%) | 0 (0%) | 0 (0%) | 0 (0%) | 0 (0%) |
| Lower extremity (knee joint) | 5 | 1 (5%) | 1 (5%) | 1 (5%) | 1 (5%) | 1 (5%) | 1 (5%) |
| | 4 | 3 (16%) | 3 (15%) | 3 (16%) | 3 (15%) | 3 (16%) | 3 (14%) |
| | 3 | 12 (63%) | 13 (65%) | 12 (63%) | 13 (65%) | 12 (63%) | 14 (67%) |
| | 2 | 3 (16%) | 3 (15%) | 3 (16%) | 3 (15%) | 3 (16%) | 3 (14%) |
| | 1 | 0 (0%) | 0 (0%) | 0 (0%) | 0 (0%) | 0 (0%) | 0 (0%) |
| | 0 | 0 (0%) | 0 (0%) | 0 (0%) | 0 (0%) | 0 (0%) | 0 (0%) |
| | Not evaluated/unknown | 0 (0%) | 0 (0%) | 0 (0%) | 0 (0%) | 0 (0%) | 0 (0%) |
| Lower extremity (ankle joint) | 5 | 0 (0%) | 1 (5%) | 0 (0%) | 1 (5%) | 0 (0%) | 1 (5%) |
| | 4 | 2 (11%) | 0 (0%) | 2 (11%) | 0 (0%) | 2 (11%) | 0 (0%) |
| | 3 | 3 (16%) | 3 (15%) | 3 (16%) | 3 (15%) | 3 (16%) | 3 (14%) |
| | 2 | 5 (26%) | 6 (30%) | 5 (26%) | 6 (30%) | 5 (26%) | 7 (33%) |
| | 1 | 6 (32%) | 7 (35%) | 6 (32%) | 7 (35%) | 6 (32%) | 7 (33%) |
| | 0 | 3 (16%) | 3 (15%) | 3 (16%) | 3 (15%) | 3 (16%) | 3 (14%) |
| | Not evaluated/unknown | 0 (0%) | 0 (0%) | 0 (0%) | 0 (0%) | 0 (0%) | 0 (0%) |

Motor Activity Log-14

**Table S11** Motor Activity Log (MAL) Amount of Use(FAS)

| Characteristics items | Statistics | REAL | | CTRL | | Group difference | |
|---|---|---|---|---|---|---|---|
| | | Measurement | Gain | Measurement | Gain | Measurement | Gain |
| Number of cases | | 19 | | 20 | | - | |
| Before-intervention | Cases | 19 | | 20 | | | |
| | Mean± S D | 0.52 ± 0.45 | | 0.71 ± 0.85 | | | |
| | Median | 0.38 | | 0.50 | | | |
| | Min. - Max. | 0.0 - 1.8 | | 0.0 - 3.6 | | | |
| After-intervention | Cases | 19 | 19 | 20 | 20 | | |
| | Mean± S D | 0.69 ± 0.51 | 0.17 ± 0.30 | 0.77 ± 0.96 | 0.06 ± 0.39 | -0.08 ± 0.77 | 0.11 ± 0.35 |
| | Median | 0.71 | 0.07 | 0.54 | 0.00 | | |
| | Min. - Max. | 0.0 - 1.9 | -0.3 - 0.9 | 0.0 - 4.1 | -0.9 - 0.7 | | |
| | LS Mean | | 0.17 | | 0.06 | | 0.11 |
| | 95% CI | | (0.01, 0.33) | | (-0.10, 0.22) | | (-0.12, 0.34) |
| | *p*-value | | | | | | 0.328 |
| Follow-up [LOCF] | Cases | 19 | 19 | 20 | 20 | | |
| | Mean± S D | 0.65 ± 0.53 | 0.14 ± 0.29 | 0.78 ± 0.92 | 0.07 ± 0.43 | -0.13 ± 0.76 | 0.07 ± 0.37 |
| | Median | 0.46 | 0.07 | 0.57 | 0.00 | | |
| | Min. - Max. | 0.0 - 1.9 | -0.4 - 0.9 | 0.0 - 3.9 | -0.9 - 1.1 | | |
| | LS Mean | | 0.13 | | 0.07 | | 0.06 |
| | 95% CI | | (-0.04, 0.31) | | (-0.10, 0.25) | | (-0.19, 0.30) |
| | *p*-value | | | | | | 0.638 |
| Follow-up [Complete Case] | Cases | 19 | 19 | 20 | 20 | | |
| | Mean± S D | 0.65 ± 0.53 | 0.14 ± 0.29 | 0.78 ± 0.92 | 0.07 ± 0.43 | -0.13 ± 0.76 | 0.07 ± 0.37 |
| | Median | 0.46 | 0.07 | 0.57 | 0.00 | | |
| | Min. - Max. | 0.0 - 1.9 | -0.4 - 0.9 | 0.0 - 3.9 | -0.9 - 1.1 | | |
| | LS Mean | | 0.13 | | 0.07 | | 0.06 |
| | 95% CI | | (-0.04, 0.31) | | (-0.10, 0.25) | | (-0.19, 0.30) |
| | *p*-value | | | | | | 0.638 |

**Table S12** Motor Activity Log (MAL) Quality of Movement(FAS)

| Characteristics items | Statistics | REAL | | CTRL | | Group difference | |
|---|---|---|---|---|---|---|---|
| | | Measurement | Gain | Measurement | Gain | Measurement | Gain |
| Number of cases | | 19 | | 20 | | - | |
| Before-intervention | Cases | 19 | | 20 | | | |
| | Mean±SD | 0.50 ± 0.45 | | 0.63 ± 0.71 | | | |
| | Median | 0.38 | | 0.43 | | | |
| | Min. - Max. | 0.0 - 1.5 | | 0.0 - 2.9 | | | |
| After-intervention | Cases | 19 | 19 | 20 | 20 | | |
| | Mean±SD | 0.53 ± 0.34 | 0.03 ± 0.27 | 0.69 ± 0.82 | 0.06 ± 0.36 | -0.15 ± 0.63 | -0.03 ± 0.32 |
| | Median | 0.42 | 0.03 | 0.46 | 0.00 | | |
| | Min. - Max. | 0.0 - 1.1 | -0.5 - 0.6 | 0.0 - 3.5 | -0.9 - 0.9 | | |
| | LS Mean | | 0.02 | | 0.07 | | -0.04 |
| | 95% CI | | (-0.13, 0.17) | | (-0.08, 0.21) | | (-0.25, 0.17) |
| | *p*-value | | | | | | 0.680 |
| Follow-up [LOCF] | Cases | 19 | 19 | 20 | 20 | | |
| | Mean±SD | 0.53 ± 0.44 | 0.02 ± 0.34 | 0.67 ± 0.74 | 0.04 ± 0.34 | -0.14 ± 0.61 | -0.02 ± 0.34 |
| | Median | 0.43 | 0.00 | 0.57 | 0.00 | | |
| | Min. - Max. | 0.0 - 1.6 | -0.9 - 0.9 | 0.0 - 3.0 | -0.9 - 0.9 | | |
| | LS Mean | | 0.02 | | 0.05 | | -0.03 |
| | 95% CI | | (-0.14, 0.17) | | (-0.10, 0.20) | | (-0.25, 0.19) |
| | *p*-value | | | | | | 0.759 |
| Follow-up [Complete Case] | Cases | 19 | 19 | 20 | 20 | | |
| | Mean±SD | 0.53 ± 0.44 | 0.02 ± 0.34 | 0.67 ± 0.74 | 0.04 ± 0.34 | -0.14 ± 0.61 | -0.02 ± 0.34 |
| | Median | 0.43 | 0.00 | 0.57 | 0.00 | | |
| | Min. - Max. | 0.0 - 1.6 | -0.9 - 0.9 | 0.0 - 3.0 | -0.9 - 0.9 | | |
| | LS Mean | | 0.02 | | 0.05 | | -0.03 |
| | 95% CI | | (-0.14, 0.17) | | (-0.10, 0.20) | | (-0.25, 0.19) |
| | *p*-value | | | | | | 0.759 |

Goal Attainment Scaling(GAS)

**Table S13** Goal Attainment Scaling(GAS) (FAS)

| Characteristics items | Categories | REAL | CTRL | *p*-value |
|---|---|---|---|---|
| Number of cases | | 19 | 20 | — |
| Before-intervention | 4 | 0 (0.0) | 0 (0.0) | — |
| | 3 | 0 (0.0) | 0 (0.0) | |
| | 2 | 0 (0.0) | 0 (0.0) | |
| | 1 | 0 (0.0) | 0 (0.0) | |
| | 0 | 0 (0.0) | 0 (0.0) | |
| | -1 | 0 (0.0) | 0 (0.0) | |
| | -2 | 2 (10.5) | 3 (15.0) | |
| | -3 | 3 (15.8) | 5 (25.0) | |
| | -4 | 14 (73.7) | 12 (60.0) | |
| After-intervention | 4 | 0 (0.0) | 0 (0.0) | 0.256 |
| | 3 | 0 (0.0) | 0 (0.0) | |
| | 2 | 0 (0.0) | 1 (5.0) | |
| | 1 | 0 (0.0) | 0 (0.0) | |
| | 0 | 3 (15.8) | 1 (5.0) | |
| | -1 | 1 (5.3) | 0 (0.0) | |
| | -2 | 6 (31.6) | 4 (20.0) | |
| | -3 | 4 (21.1) | 7 (35.0) | |
| | -4 | 5 (26.3) | 7 (35.0) | |
| Follow-up [LOCF] | 4 | 0 (0.0) | 1 (5.0) | 0.075 |
| | 3 | 0 (0.0) | 0 (0.0) | |
| | 2 | 0 (0.0) | 1 (5.0) | |
| | 1 | 0 (0.0) | 0 (0.0) | |
| | 0 | 1 (5.3) | 0 (0.0) | |
| | -1 | 6 (31.6) | 0 (0.0) | |
| | -2 | 5 (26.3) | 4 (20.0) | |
| | -3 | 4 (21.1) | 9 (45.0) | |
| | -4 | 3 (15.8) | 5 (25.0) | |
| Follow-up [Complete Case] | 4 | 0 (0.0) | 1 (5.0) | 0.075 |
| | 3 | 0 (0.0) | 0 (0.0) | |
| | 2 | 0 (0.0) | 1 (5.0) | |
| | 1 | 0 (0.0) | 0 (0.0) | |
| | 0 | 1 (5.3) | 0 (0.0) | |
| | -1 | 6 (31.6) | 0 (0.0) | |
| | -2 | 5 (26.3) | 4 (20.0) | |
| | -3 | 4 (21.1) | 9 (45.0) | |
| | -4 | 3 (15.8) | 5 (25.0) | |

ARAT

**Table S14** ARAT (action research arm test)(FAS)

| Characteristics items | Statistics | REAL | | CTRL | | Group difference | |
|---|---|---|---|---|---|---|---|
| | | Measurement | Difference | Measurement | Difference | Measurement | Difference |
| | Number of cases | 19 | | 20 | | - | |
| Before-intervention | Cases | 19 | | 20 | | | |
| | Mean± S D | 4.6 ± 4.4 | | 5.1 ± 5.7 | | | |
| | Median | 3.0 | | 3.5 | | | |
| | Min. - Max. | 0 - 18 | | 0 - 26 | | | |
| After-intervention | Cases | 19 | 19 | 20 | 20 | | |
| | Mean± S D | 5.5 ± 4.4 | 0.9 ± 1.7 | 6.3 ± 6.6 | 1.2 ± 2.0 | -0.7 ± 5.6 | -0.3 ± 1.9 |
| | Median | 5.0 | 0.0 | 4.5 | 0.0 | | |
| | Min. - Max. | 0 - 18 | -1 - 5 | 0 - 30 | 0 - 8 | | |
| | LS Mean | | 1.0 | | 1.2 | | -0.2 |
| | 95% CI | | (0.1, 1.8) | | (0.3, 2.1) | | (-1.5, 1.0) |
| | p-value | | | | | | 0.700 |
| Follow-up [LOCF] | Cases | 19 | 19 | 20 | 20 | | |
| | Mean± S D | 7.1 ± 5.9 | 2.5 ± 3.4 | 6.5 ± 6.0 | 1.4 ± 2.0 | 0.6 ± 6.0 | 1.1 ± 2.8 |
| | Median | 6.0 | 2.0 | 5.0 | 1.0 | | |
| | Min. - Max. | 0 - 23 | -2 - 12 | 0 - 27 | 0 - 8 | | |
| | LS Mean | | 2.5 | | 1.4 | | 1.1 |
| | 95% CI | | (1.2, 3.8) | | (0.1, 2.7) | | (-0.8, 2.9) |
| | p-value | | | | | | 0.242 |
| Follow-up [Complete Case] | Cases | 19 | 19 | 20 | 20 | | |
| | Mean± S D | 7.1 ± 5.9 | 2.5 ± 3.4 | 6.5 ± 6.0 | 1.4 ± 2.0 | 0.6 ± 6.0 | 1.1 ± 2.8 |
| | Median | 6.0 | 2.0 | 5.0 | 1.0 | | |
| | Min. - Max. | 0 - 23 | -2 - 12 | 0 - 27 | 0 - 8 | | |
| | LS Mean | | 2.5 | | 1.4 | | 1.1 |
| | 95% CI | | (1.2, 3.8) | | (0.1, 2.7) | | (-0.8, 2.9) |
| | p-value | | | | | | 0.242 |

Barthel Index

**Table S15** Barthel Index (FAS)

| Characteristics items | Statistics | REAL | | CTRL | | Group difference | |
|---|---|---|---|---|---|---|---|
| | | Measurement | Gain | Measurement | Gain | Measurement | Gain |
| | Number of cases | 19 | | 20 | | - | |
| Before-intervention | Cases | 19 | | 20 | | | |
| | Mean± S D | 97.6 ± 7.1 | | 98.5 ± 4.0 | | | |
| | Median | 100.0 | | 100.0 | | | |
| | Min. - Max. | 70 - 100 | | 85 - 100 | | | |
| After-intervention [Complete Case] | Cases | 19 | 19 | 20 | 20 | | |
| | Mean± S D | 98.4 ± 3.7 | 0.8 ± 4.2 | 97.8 ± 4.7 | -0.8 ± 4.1 | 0.7 ± 4.3 | 1.5 ± 4.1 |
| | Median | 100.0 | 0.0 | 100.0 | 0.0 | | |
| | Min. - Max. | 85 - 100 | -5 - 15 | 85 - 100 | -10 - 10 | | |
| | LS Mean | | 0.6 | | -0.5 | | 1.1 |
| | 95% CI | | (-0.9, 2.0) | | (-2.0, 0.9) | | (-0.9, 3.1) |
| | *p*-value | | | | | | 0.267 |

SS-QOL (stroke specific quality of life)(FAS)

**Table S16** SS-QOL (stroke specific quality of life)(FAS)

| Characteristics items | Statistics | REAL | | CTRL | | Group difference | |
|---|---|---|---|---|---|---|---|
| | | Measurement | Gain | Measurement | Gain | Measurement | Gain |
| Number of cases | | 19 | | 20 | | - | |
| Before-intervention | Cases | 19 | | 19 | | | |
| | Mean± S D | 180.2 ± 28.0 | | 184.3 ± 32.8 | | | |
| | Median | 177.0 | | 189.0 | | | |
| | Min. - Max. | 134 - 234 | | 131 - 234 | | | |
| Follow-up [Complete Case] | Cases | 19 | 19 | 20 | 19 | | |
| | Mean± S D | 189.5 ± 30.6 | 9.3 ± 18.9 | 198.9 ± 30.9 | 13.9 ± 21.2 | -9.4 ± 30.8 | -4.6 ± 20.1 |
| | Median | 186.0 | 11.0 | 201.0 | 4.0 | | |
| | Min. - Max. | 140 - 245 | -16 - 62 | 106 - 238 | -33 - 59 | | |
| | LS Mean | | 8.9 | | 14.3 | | -5.4 |
| | 95% CI | | (-0.1, 18.0) | | (5.2, 23.4) | | (-18.2, 7.4) |
| | *p*-value | | | | | | 0.399 |

Modified Ashworth Scale

**Table S17** MAS (modified Ashworth scale) (FAS)

| Characteristics items | Statistics | REAL | | CTRL | | Group difference | |
|---|---|---|---|---|---|---|---|
| | | Measurement | Gain | Measurement | Gain | Measurement | Gain |
| Number of cases | | 19 | | 20 | | - | |
| Before-intervention | Cases | 19 | | 20 | | | |
| | Mean± S D | 6.7 ± 1.8 | | 6.9 ± 2.3 | | | |
| | Median | 6.0 | | 6.0 | | | |
| | Min. - Max. | 3 - 9 | | 3 - 12 | | | |
| After-intervention | Cases | 19 | 19 | 20 | 20 | | |
| | Mean± S D | 6.2 ± 1.8 | -0.5 ± 2.0 | 6.1 ± 2.4 | -0.8 ± 2.0 | 0.2 ± 2.2 | 0.3 ± 2.0 |
| | Median | 6.0 | 0.0 | 6.0 | 0.0 | | |
| | Min. - Max. | 3 - 10 | -5 - 4 | 1 - 10 | -5 - 2 | | |
| | LS Mean | | -0.5 | | -0.8 | | 0.3 |
| | 95% CI | | (-1.4, 0.3) | | (-1.6, 0.1) | | (-0.9, 1.4) |
| | *p*-value | | | | | | 0.668 |
| Follow-up [LOCF] | Cases | 19 | 19 | 20 | 20 | | |
| | Mean± S D | 6.4 ± 1.9 | -0.3 ± 1.4 | 7.1 ± 2.2 | 0.3 ± 1.5 | -0.7 ± 2.1 | -0.5 ± 1.5 |
| | Median | 6.0 | 0.0 | 7.0 | 0.0 | | |
| | Min. - Max. | 3 - 9 | -3 - 3 | 3 - 12 | -3 - 3 | | |
| | LS Mean | | -0.3 | | 0.3 | | -0.6 |
| | 95% CI | | (-0.9, 0.4) | | (-0.4, 0.9) | | (-1.5, 0.3) |
| | *p*-value | | | | | | 0.216 |
| Follow-up [Complete Case] | Cases | 19 | 19 | 20 | 20 | | |
| | Mean± S D | 6.4 ± 1.9 | -0.3 ± 1.4 | 7.1 ± 2.2 | 0.3 ± 1.5 | -0.7 ± 2.1 | -0.5 ± 1.5 |
| | Median | 6.0 | 0.0 | 7.0 | 0.0 | | |
| | Min. - Max. | 3 - 9 | -3 - 3 | 3 - 12 | -3 - 3 | | |
| | LS Mean | | -0.3 | | 0.3 | | -0.6 |
| | 95% CI | | (-0.9, 0.4) | | (-0.4, 0.9) | | (-1.5, 0.3) |
| | *p*-value | | | | | | 0.216 |

#### Raw Clinical Data

**Table S18** Clinical assessment scores

| Patints | Group | Time points and measure | | FMA | | | | | Barthel Index | MAL | | SIAS | | | | | GAS | | MAS | | | SSQOL | ARAT |
|---|---|---|---|---|---|---|---|---|---|---|---|---|---|---|---|---|---|---|---|---|---|---|---|
| | | | | Total | A (Soulder-elbow-forearm) | B (Wrist) | C (Finger) | D (Cordination and speed) | | AOU | QOM | Upper extremity (proximal) | Fingers | Lower extermity (hip joint) | Lower extermity (knee joint) | Lower extermity (ankle joint) | item1 | item2 | Fingers | Wrist | Elbow | | |
| 1 | REAL | Before-intervention | 04/2017 | 15 | 10 | 0 | 5 | 0 | - | 0.2 | 0.2 | 2 | 1a | 3 | 3 | 1 | -2 | -2 | 2 | 1 | 2 | 153 | - |
| | | | 04/2017 | - | - | - | - | - | 70 | - | - | - | - | - | - | - | - | - | - | - | - | - | 3 |
| | | After-intervention | 04/2017 | 19 | 13 | 2 | 4 | 0 | - | 0.8 | 0.3 | - | - | - | - | - | - | - | 2 | 1+ | 2 | - | 3 |
| | | | 04/2017 | - | - | - | - | - | - | - | - | 2 | 1a | 3 | 3 | 1 | -2 | -2 | - | - | - | - | - |
| | | Follow-up | 05/2017 | 28 | 18 | 3 | 7 | 0 | 85 | 0.4 | 0.5 | 3 | 1a | 3 | 3 | 1 | -1 | -2 | 1 | 1+ | 1+ | 140 | 5 |
| 2 | CTRL | Before-intervention | 05/2017 | 20 | 14 | 0 | 6 | 0 | - | 1.0 | 0.9 | 2 | 1a | 4 | 3 | 1 | -2 | -1 | 1+ | 2 | 2 | 215 | - |
| | | | 05/2017 | - | - | - | - | - | 100 | - | - | - | - | - | - | - | - | - | - | - | - | - | 2 |
| | | After-intervention | 05/2017 | - | - | - | - | - | - | - | - | - | - | - | - | - | - | - | - | - | - | - | 3 |
| | | | 05/2017 | 30 | 18 | 2 | 10 | 0 | - | 1.7 | 1.8 | 2 | 1a | 3 | 3 | 0 | +1 | +1 | 1+ | 3 | 2 | - | - |
| | | Follow-up | 06/2017 | 25 | 17 | 1 | 7 | 0 | 100 | 2.1 | 1.8 | 3 | 1a | 4 | 3 | 1 | +2 | +2 | 1+ | 2 | 1+ | 228 | 2 |
| 3 | REAL | Before-intervention | 05/2017 | 14 | 9 | 0 | 5 | 0 | - | 0.0 | 0.0 | 3 | 1a | 2 | 2 | 1 | -2 | -2 | 2 | 2 | 2 | 200 | - |
| | | | 06/2017 | - | - | - | - | - | 90 | - | - | - | - | - | - | - | - | - | - | - | - | - | 4 |
| | | After-intervention | 06/2017 | 24 | 16 | 0 | 8 | 0 | - | 0.2 | 0.3 | 3 | 1a | 2 | 2 | 2 | -2 | -2 | 1+ | 1+ | 1 | - | 6 |
| | | Follow-up | 07/2017 | 24 | 13 | 3 | 8 | 0 | 95 | 0.3 | 0.1 | 3 | 1a | 3 | 3 | 0 | -1 | 0 | 1+ | 1+ | 2 | 184 | 6 |
| 4 | REAL | Before-intervention | 07/2017 | 10 | 9 | 0 | 1 | 0 | 100 | 0.5 | 0.4 | 2 | 1a | 4 | 4 | 2 | -2 | -2 | 3 | 2 | 1+ | 187 | 0 |
| | | After-intervention | 08/2017 | 10 | 9 | 0 | 1 | 0 | - | 0.7 | 0.6 | 2 | 1a | 4 | 4 | 2 | 0 | -1 | 3 | 1+ | 1+ | - | 0 |
| | | Follow-up | 09/2017 | 10 | 9 | 0 | 1 | 0 | 100 | 0.6 | 0.6 | 2 | 1a | 4 | 4 | 2 | 0 | -2 | 3 | 1+ | 2 | 210 | 0 |
| 5 | CTRL | Before-intervention | 08/2017 | - | - | - | - | - | - | - | - | 2 | 1a | 3 | 3 | 1 | -2 | -2 | - | - | - | 224 | - |
| | | | 08/2017 | 9 | 8 | 0 | 1 | 0 | 100 | 0.0 | 0.1 | - | - | - | - | - | - | - | 2 | 2 | 1+ | - | 0 |
| | | After-intervention | 08/2017 | - | - | - | - | - | - | - | - | 3 | 1a | 3 | 3 | 1 | -2 | -1 | - | - | - | - | 1 |
| | | | 08/2017 | 14 | 12 | 0 | 2 | 0 | - | 0.4 | 0.4 | - | - | - | - | - | - | - | 2 | 2 | 0 | - | - |
| | | Follow-up | 09/2017 | 13 | 11 | 0 | 2 | 0 | 100 | 0.4 | 0.2 | 3 | 1a | 3 | 3 | 1 | -2 | -1 | 1+ | 2 | 1+ | 227 | 4 |
| 6 | REAL | Before-intervention | 08/2017 | 23 | 17 | 3 | 3 | 0 | 100 | 1.8 | 1.5 | 3 | 1a | 5 | 5 | 0 | -2 | -2 | 1+ | 2 | 1 | 134 | 7 |
| | | After-intervention | 09/2017 | 29 | 21 | 4 | 4 | 0 | - | 1.9 | 1.1 | 3 | 1a | 5 | 5 | 0 | -2 | -2 | 1+ | 2 | 1 | - | 7 |
| | | Follow-up | 10/2017 | 31 | 23 | 4 | 4 | 0 | 100 | 1.9 | 1.3 | 3 | 1a | 5 | 5 | 0 | -2 | -2 | 1+ | 2 | 1 | 146 | 7 |
| 7 | CTRL | Before-intervention | 08/2017 | 15 | 9 | 0 | 6 | 0 | - | 0.9 | 0.7 | 2 | 1a | 3 | 3 | 1 | -2 | -2 | 1 | 1+ | 2 | - | - |
| | | | 08/2017 | - | - | - | - | - | 100 | - | - | - | - | - | - | - | - | - | - | - | - | - | 3 |
| | | | 08/2017 | - | - | - | - | - | - | - | - | - | - | - | - | - | - | - | - | - | - | 152 | - |
| | | After-intervention | 09/2017 | - | - | - | - | - | - | - | - | 2 | 1a | 4 | 3 | 1 | -2 | -1 | - | - | - | - | 3 |
| | | | 09/2017 | 13 | 8 | 0 | 5 | 0 | - | 0.5 | 0.6 | - | - | - | - | - | - | - | 0 | 0 | 1 | - | - |
| | | Follow-up | 10/2017 | 20 | 12 | 0 | 8 | 0 | 100 | 0.6 | 0.6 | 2 | 1a | 3 | 3 | 0 | -2 | -1 | 1 | 1+ | 1+ | 183 | 3 |
| 8 | REAL | Before-intervention | 09/2017 | 33 | 26 | 0 | 7 | 0 | - | 0.3 | 0.2 | 3 | 1a | 4 | 4 | 4 | - | - | 1 | 2 | 1 | - | - |
| | | | 09/2017 | - | - | - | - | - | 100 | - | - | - | - | - | - | - | -2 | -2 | - | - | - | 168 | 7 |
| | | After-intervention | 10/2017 | 38 | 30 | 0 | 8 | 0 | - | 0.1 | 0.1 | 4 | 1b | 4 | 4 | 4 | -2 | 0 | 1+ | 1 | 1+ | - | 7 |
| | | Follow-up | 11/2017 | 43 | 29 | 4 | 10 | 0 | 95 | 0.2 | 0.1 | 4 | 1b | 4 | 4 | 4 | -2 | +1 | 1 | 2 | 1+ | 184 | 15 |
| 9 | CTRL | Before-intervention | 10/2017 | 20 | 19 | 0 | 1 | 0 | 100 | 0.1 | 0.1 | 3 | 1a | 3 | 3 | 1 | -1 | -2 | 1 | 1+ | 1+ | - | 11 |
| | | | 10/2017 | - | - | - | - | - | - | - | - | - | - | - | - | - | - | - | - | - | - | 201 | - |
| | | After-intervention | 10/2017 | 23 | 22 | 0 | 1 | 0 | - | 0.6 | 0.4 | 3 | 1a | 3 | 3 | 1 | -1 | -2 | 0 | 0 | 1 | - | 13 |
| | | Follow-up | 11/2017 | 22 | 21 | 0 | 1 | 0 | 100 | 0.7 | 0.6 | 3 | 1a | 3 | 3 | 1 | -1 | -2 | 0 | 1+ | 1 | 203 | 14 |
| 10 | CTRL | Before-intervention | 11/2017 | 13 | 12 | 0 | 1 | 0 | - | 0.1 | 0.1 | 2 | 1a | 3 | 4 | 3 | -1 | -2 | 0 | 1+ | 1 | 189 | - |
| | | | 11/2017 | - | - | - | - | - | 100 | - | - | - | - | - | - | - | - | - | - | - | - | - | 1 |
| | | After-intervention | 11/2017 | - | - | - | - | - | - | - | - | - | - | - | - | - | - | - | - | - | - | - | 1 |
| | | | 11/2017 | 15 | 13 | 0 | 2 | 0 | - | 0.1 | 0.2 | 3 | 1a | 3 | 4 | 3 | -2 | -2 | 1 | 1+ | 1+ | - | - |
| | | Follow-up | 12/2017 | 19 | 15 | 0 | 4 | 0 | 90 | 0.0 | 0.0 | 3 | 1a | 3 | 4 | 3 | -1 | -1 | 0 | 1+ | 1+ | 187 | 1 |
| 11 | CTRL | Before-intervention | 11/2017 | 18 | 17 | 0 | 1 | 0 | 100 | 0.3 | 0.3 | 3 | 1a | 4 | 2 | 0 | -2 | -2 | 3 | 3 | 3 | 202 | 3 |
| | | After-intervention | 12/2017 | 18 | 17 | 0 | 1 | 0 | - | 0.4 | 0.4 | 3 | 1a | 4 | 2 | 0 | -1 | -1 | 2 | 2 | 3 | - | 3 |
| | | Follow-up | 01/2018 | 18 | 17 | 0 | 1 | 0 | 100 | 0.4 | 0.3 | 3 | 1a | 4 | 2 | 0 | -2 | -2 | 3 | 3 | 3 | 205 | 3 |
| 12 | REAL | Before-intervention | 12/2017 | - | - | - | - | - | - | - | - | 2 | 1a | 3 | 3 | 3 | -2 | -2 | - | - | - | 155 | - |
| | | | 12/2017 | 16 | 10 | 0 | 6 | 0 | 100 | 0.6 | 0.6 | - | - | - | - | - | - | - | 1+ | 1+ | 1+ | - | 1 |
| | | After-intervention | 12/2017 | 18 | 11 | 0 | 7 | 0 | - | 1.1 | 1.0 | 2 | 1a | 3 | 3 | 3 | 0 | 0 | 1+ | 2 | 2 | - | 2 |
| | | Follow-up | 02/2018 | 17 | 12 | 0 | 5 | 0 | 100 | 1.0 | 0.9 | 2 | 1a | 3 | 3 | 3 | 0 | -1 | 2 | 1 | 1+ | 154 | 2 |
| 13 | REAL | Before-intervention | 12/2017 | - | - | - | - | - | - | - | - | 2 | 1a | 3 | 2 | 0 | -2 | -2 | - | - | - | - | - |
| | | | 12/2017 | 19 | 8 | 4 | 7 | 0 | 100 | 0.2 | 0.2 | - | - | - | - | - | - | - | 1+ | 1+ | 2 | 199 | 3 |
| | | After-intervention | 12/2017 | 18 | 9 | 2 | 7 | 0 | - | 0.4 | 0.3 | 3 | 1b | 3 | 3 | 1 | -2 | -2 | 2 | 1+ | 2 | - | 7 |
| | | Follow-up | 01/2018 | 16 | 8 | 0 | 8 | 0 | 100 | 0.2 | 0.1 | 3 | 1a | 3 | 3 | 1 | -1 | -1 | 1+ | 2 | 2 | 208 | 6 |
| 14 | CTRL | Before-intervention | 12/2017 | 29 | 28 | 0 | 1 | 0 | 100 | 0.6 | 0.5 | 4 | 1a | 4 | 4 | 3 | - | - | 2 | 1+ | 1+ | 178 | 3 |
| | | | 12/2017 | - | - | - | - | - | - | - | - | - | - | - | - | - | -1 | -1 | - | - | - | - | - |
| | | After-intervention | 12/2017 | 29 | 28 | 0 | 1 | 0 | - | 0.6 | 0.5 | 4 | 1a | 4 | 4 | 3 | -1 | -1 | 2 | 1+ | 1+ | - | 3 |
| | | Follow-up | 01/2018 | 30 | 29 | 0 | 1 | 0 | 100 | 0.6 | 0.5 | 4 | 1a | 4 | 4 | 3 | -1 | -1 | 2 | 1+ | 1+ | 178 | 3 |
| 15 | CTRL | Before-intervention | 01/2018 | 19 | 15 | 0 | 4 | 0 | - | 1.0 | 0.9 | 2 | 1a | 3 | 3 | 2 | -2 | -2 | 1+ | 2 | 1 | - | - |
| | | | 01/2018 | - | - | - | - | - | 100 | - | - | - | - | - | - | - | - | - | - | - | - | - | 4 |
| | | After-intervention | 01/2018 | 20 | 19 | 0 | 1 | 0 | - | 1.5 | 1.2 | - | - | - | - | - | - | - | 1+ | 2 | 1+ | 209 | - |
| | | | 01/2018 | - | - | - | - | - | - | - | - | 2 | 1a | 3 | 3 | 2 | -2 | -2 | - | - | - | - | 4 |
| | | Follow-up | 02/2018 | 19 | 16 | 0 | 3 | 0 | 90 | 1.5 | 1.2 | 3 | 1a | 3 | 3 | 1 | -1 | -2 | 1 | 2 | 1 | 212 | 4 |
| 16 | CTRL | Before-intervention | 01/2018 | 22 | 19 | 0 | 3 | 0 | - | - | - | 3 | 1a | 4 | 3 | 3 | -1 | -2 | - | - | - | 200 | - |
| | | | 01/2018 | - | - | - | - | - | 100 | 1.2 | 1.3 | - | - | - | - | - | - | - | 2 | 2 | 1+ | - | 5 |
| | | After-intervention | 02/2018 | 20 | 17 | 0 | 3 | 0 | - | 1.2 | 1.4 | 3 | 1a | 4 | 3 | 3 | -1 | -2 | 1 | 2 | 1+ | - | 5 |
| | | Follow-up | 03/2018 | 26 | 23 | 0 | 3 | 0 | 100 | 1.4 | 1.5 | 3 | 1a | 4 | 3 | 3 | -1 | -2 | 2 | 3 | 2 | 199 | 6 |
| 17 | REAL | Before-intervention | 02/2018 | - | - | - | - | - | - | - | - | 3 | 1a | 4 | 3 | 3 | -2 | -2 | - | - | - | - | - |
| | | | 02/2018 | 26 | 16 | 5 | 5 | 0 | 100 | 0.6 | 0.8 | - | - | - | - | - | - | - | 2 | 2 | 2 | 177 | 1 |
| | | After-intervention | 02/2018 | 29 | 19 | 4 | 6 | 0 | - | 1.3 | 0.9 | 3 | 1b | 4 | 3 | 3 | -1 | +1 | 2 | 1+ | 2 | - | 1 |
| | | Follow-up | 03/2018 | 27 | 17 | 4 | 6 | 0 | 100 | 1.6 | 1.6 | 3 | 1a | 4 | 3 | 3 | -1 | +1 | 2 | 2 | 2 | 215 | 3 |
| 18 | CTRL | Before-intervention | 01/2018 | 28 | 21 | 2 | 5 | 0 | - | 1.3 | 1.0 | 3 | 1a | 4 | 3 | 1 | -2 | -2 | 1 | 1+ | 2 | 180 | - |
| | | | 01/2018 | - | - | - | - | - | 100 | - | - | - | - | - | - | - | - | - | - | - | - | - | 4 |
| | | After-intervention | 02/2018 | 22 | 15 | 2 | 5 | 0 | - | 0.6 | 0.6 | - | - | - | - | - | - | - | 1+ | 1+ | 2 | - | - |
| | | | 02/2018 | - | - | - | - | - | - | - | - | 3 | 1a | 4 | 3 | 1 | -2 | -2 | - | - | - | - | 7 |
| | | Follow-up | 03/2018 | 24 | 16 | 2 | 6 | 0 | 100 | 0.6 | 0.6 | 3 | 1a | 4 | 3 | 1 | -2 | -1 | 1+ | 2 | 2 | 228 | 8 |
| 19 | REAL | Before-intervention | 02/2018 | 20 | 15 | 0 | 5 | 0 | - | 0.2 | 0.3 | 3 | 1a | 3 | 3 | 1 | -2 | -2 | 2 | 2 | 2 | - | - |
| | | | 02/2018 | - | - | - | - | - | 100 | - | - | - | - | - | - | - | - | - | - | - | - | 183 | 2 |
| | | After-intervention | 03/2018 | 21 | 16 | 0 | 5 | 0 | - | 1.1 | 0.9 | 3 | 1b | 3 | 3 | 1 | 0 | 0 | 2 | 1+ | 2 | - | 2 |
| | | Follow-up | 04/2018 | 24 | 17 | 2 | 5 | 0 | 100 | 0.9 | 0.6 | 3 | 1b | 3 | 3 | 2 | 0 | -1 | 2 | 1+ | 2 | 188 | 9 |
| 20 | CTRL | Before-intervention | 03/2018 | - | - | - | - | - | 100 | - | - | 2 | 1a | 3 | 3 | 1 | -2 | -2 | - | - | - | 226 | 0 |
| | | | 03/2018 | 12 | 10 | 0 | 2 | 0 | - | 0.0 | 0.0 | - | - | - | - | - | - | - | 2 | 1 | 2 | - | - |
| | | After-intervention | 03/2018 | 14 | 11 | 0 | 3 | 0 | - | 0.0 | 0.0 | - | - | - | - | - | - | - | 1 | 1 | 1+ | - | - |
| | | | 03/2018 | - | - | - | - | - | - | - | - | 2 | 1a | 3 | 3 | 1 | -2 | -2 | - | - | - | - | 0 |
| | | Follow-up | 04/2018 | 14 | 11 | 0 | 3 | 0 | 100 | 0.0 | 0.0 | 2 | 1a | 3 | 3 | 1 | -2 | -2 | 1 | 1+ | 3 | 228 | 0 |
| 21 | CTRL | Before-intervention | 02/2018 | 21 | 18 | 0 | 3 | 0 | 85 | 0.2 | 0.2 | 3 | 1a | 3 | 2 | 2 | -2 | -2 | 2 | 2 | 2 | 139 | 4 |
| | | After-intervention | 03/2018 | 21 | 18 | 0 | 3 | 0 | - | 0.2 | 0.2 | 3 | 1a | 3 | 3 | 2 | -1 | -2 | 1+ | 1+ | 1+ | - | 5 |
| | | Follow-up | 04/2018 | 21 | 18 | 0 | 3 | 0 | 85 | 0.2 | 0.2 | 3 | 1a | 3 | 3 | 2 | -1 | -2 | 2 | 2 | 2 | 106 | 5 |
| 22 | CTRL | Before-intervention | 02/2018 | - | - | - | - | - | 100 | - | - | 2 | 1a | 5 | 5 | 5 | -2 | -2 | - | - | - | 209 | 2 |
| | | | 02/2018 | 15 | 13 | 0 | 2 | 0 | - | 0.0 | 0.0 | - | - | - | - | - | - | - | 1 | 1+ | 1+ | - | - |
| | | After-intervention | 03/2018 | 18 | 14 | 1 | 3 | 0 | - | 0.0 | 0.0 | - | - | - | - | - | - | - | 1 | 1+ | 1+ | - | - |
| | | | 03/2018 | - | - | - | - | - | - | - | - | 3 | 1a | 5 | 5 | 5 | -1 | -2 | - | - | - | - | 10 |
| | | Follow-up | 04/2018 | - | - | - | - | - | 100 | - | - | 2 | 1a | 5 | 5 | 5 | -1 | -2 | - | - | - | 234 | 10 |
| | | | 04/2018 | 22 | 15 | 4 | 3 | 0 | - | 0.0 | 0.0 | - | - | - | - | - | - | - | 1 | 2 | 2 | - | - |
| 23 | REAL | Before-intervention | 03/2018 | 23 | 20 | 0 | 3 | 0 | - | 0.0 | 0.0 | - | - | - | - | - | - | - | 0 | 1 | 1+ | - | - |
| | | | 03/2018 | - | - | - | - | - | 100 | - | - | 3 | 1a | 3 | 3 | 1 | -1 | -1 | - | - | - | 173 | 1 |
| | | After-intervention | 03/2018 | 25 | 22 | 0 | 3 | 0 | - | 0.0 | 0.0 | 3 | 1a | 3 | 3 | 3 | -1 | -1 | 0 | 1 | 1+ | - | 1 |
| | | Follow-up | 04/2018 | 22 | 19 | 1 | 2 | 0 | 100 | 0.0 | 0.0 | 3 | 1a | 3 | 3 | 1 | -1 | -1 | 0 | 1 | 1+ | 160 | 4 |
| 24 | REAL | Before-intervention | 03/2018 | - | - | - | - | - | - | - | - | 2 | 1a | 3 | 3 | 1 | -2 | -2 | - | - | - | - | - |
| | | | 03/2018 | 17 | 12 | 0 | 5 | 0 | 100 | 0.2 | 0.1 | - | - | - | - | - | - | - | 1+ | 1 | 2 | 164 | 3 |
| | | After-intervention | 03/2018 | 16 | 9 | 2 | 5 | 0 | - | 0.3 | 0.4 | 3 | 1a | 4 | 3 | 2 | -1 | -2 | 1 | 1+ | 2 | - | 7 |
| | | Follow-up | 04/2018 | 18 | 12 | 1 | 5 | 0 | 95 | 0.2 | 0.2 | 3 | 1a | 4 | 3 | 1 | -1 | -2 | 1+ | 1+ | 2 | 176 | 6 |
| 25 | CTRL | Before-intervention | 03/2018 | 32 | 22 | 4 | 6 | 0 | - | 1.6 | 1.6 | 3 | 1a | 4 | 3 | 1 | -1 | -1 | 0 | 2 | 1 | - | - |
| | | | 03/2018 | - | - | - | - | - | 100 | - | - | - | - | - | - | - | - | - | - | - | - | 212 | 11 |
| | | After-intervention | 03/2018 | 35 | 23 | 4 | 7 | 1 | - | 0.7 | 0.6 | 3 | 1a | 4 | 3 | 1 | -1 | -1 | 1 | 1+ | 1 | - | 11 |
| | | Follow-up | 04/2018 | 35 | 25 | 3 | 6 | 1 | 100 | 0.6 | 0.6 | 3 | 1a | 4 | 3 | 1 | -1 | -1 | 0 | 1+ | 2 | 213 | 12 |
| 26 | REAL | Before-intervention | 03/2018 | 27 | 24 | 2 | 1 | 0 | 100 | 1.0 | 1.1 | 3 | 1a | 4 | 4 | 4 | -2 | -1 | 1 | 1+ | 1+ | 206 | 6 |
| | | After-intervention | 04/2018 | 29 | 25 | 2 | 2 | 0 | - | 0.8 | 0.8 | 3 | 1a | 4 | 4 | 4 | -2 | -1 | 1 | 1+ | 1+ | - | 11 |
| | | Follow-up | 04/2018 | 28 | 26 | 0 | 2 | 0 | 100 | 1.1 | 1.1 | 3 | 1b | 4 | 4 | 4 | -1 | -1 | 1 | 1+ | 1+ | 223 | 18 |
| 27 | REAL | Before-intervention | 04/2018 | 14 | 11 | 0 | 3 | 0 | - | 0.0 | 0.0 | - | - | - | - | - | - | - | 1+ | 1+ | 1+ | - | - |
| | | | 04/2018 | - | - | - | - | - | 100 | - | - | 2 | 1a | 3 | 3 | 2 | -1 | -1 | - | - | - | 230 | 0 |
| | | After-intervention | 04/2018 | - | - | - | - | - | - | - | - | 2 | 1a | 3 | 3 | 2 | -1 | -1 | - | - | - | - | 1 |
| | | | 04/2018 | 18 | 15 | 0 | 3 | 0 | - | 0.0 | 0.0 | - | - | - | - | - | - | - | 1+ | 3 | 3 | - | - |
| | | Follow-up | 05/2018 | 19 | 16 | 0 | 3 | 0 | 100 | 0.0 | 0.0 | 2 | 1a | 3 | 3 | 2 | -1 | -1 | 1+ | 1+ | 2 | 236 | 0 |
| 28 | CTRL | Before-intervention | 03/2018 | 15 | 10 | 0 | 5 | 0 | 90 | 0.4 | 0.4 | 3 | 1a | 4 | 2 | 2 | -2 | -2 | 1 | 1+ | 2 | 159 | 3 |
| | | After-intervention | 04/2018 | - | - | - | - | - | - | - | - | - | - | - | - | - | - | - | - | - | - | - | 3 |
| | | | 04/2018 | 13 | 10 | 0 | 3 | 0 | - | 0.4 | 0.3 | 3 | 1a | 4 | 2 | 2 | -2 | -2 | 1+ | 1+ | 1+ | - | - |
| | | Follow-up | 05/2018 | 21 | 14 | 3 | 4 | 0 | 100 | 0.6 | 0.6 | 2 | 1a | 3 | 2 | 2 | -2 | -2 | 2 | 1+ | 2 | 183 | 5 |
| 29 | CTRL | Before-intervention | 04/2018 | 22 | 21 | 0 | 1 | 0 | 100 | 0.6 | 0.6 | 3 | 1a | 4 | 4 | 0 | -1 | -2 | 3 | 3 | 2 | 234 | 4 |
| | | After-intervention | 04/2018 | 23 | 22 | 0 | 1 | 0 | - | 0.6 | 0.6 | 3 | 1a | 4 | 4 | 0 | 0 | 0 | 2 | 2 | 1+ | - | 5 |
| | | Follow-up | 05/2018 | 23 | 22 | 0 | 1 | 0 | 100 | 0.7 | 0.6 | 3 | 1a | 4 | 4 | 0 | +1 | +1 | 2 | 2 | 1+ | 238 | 5 |
| 30 | REAL | Before-intervention | 04/2018 | 29 | 26 | 0 | 3 | 0 | 100 | 0.4 | 0.4 | 3 | 1a | 3 | 3 | 3 | -2 | -2 | 0 | 1+ | 2 | 192 | 18 |
| | | After-intervention | 05/2018 | 31 | 27 | 0 | 4 | 0 | - | 0.4 | 0.4 | 3 | 1a | 4 | 3 | 2 | 0 | -2 | 0 | 1+ | 2 | - | 18 |
| | | Follow-up | 06/2018 | 34 | 29 | 1 | 4 | 0 | 100 | 0.5 | 0.4 | 3 | 1a | 4 | 3 | 2 | -2 | -2 | 0 | 1 | 2 | 183 | 23 |
| 31 | REAL | Before-intervention | 05/2018 | 17 | 11 | 0 | 6 | 0 | 100 | 1.0 | 0.6 | 2 | 1a | 3 | 2 | 2 | -1 | -2 | 1+ | 1+ | 1+ | 135 | 5 |
| | | After-intervention | 05/2018 | 22 | 15 | 2 | 5 | 0 | - | 1.4 | 0.6 | 2 | 1a | 3 | 2 | 2 | -1 | -1 | 1 | 1 | 2 | - | 4 |
| | | Follow-up | 06/2018 | 24 | 17 | 2 | 5 | 0 | 100 | 1.4 | 0.6 | 2 | 1a | 3 | 2 | 2 | 0 | -1 | 1 | 1+ | 2 | 151 | 6 |
| 32 | CTRL | Before-intervention | 05/2018 | - | - | - | - | - | - | - | - | 3 | 1a | 3 | 3 | 2 | -2 | -2 | - | - | - | - | - |
| | | | 05/2018 | 27 | 18 | 4 | 5 | 0 | 100 | 1.4 | 0.8 | - | - | - | - | - | - | - | 2 | 1+ | 1 | 159 | 6 |
| | | After-intervention | 05/2018 | 29 | 22 | 2 | 5 | 0 | - | 1.7 | 1.0 | 3 | 1a | 3 | 3 | 2 | -2 | -2 | 1+ | 1+ | 1+ | - | 9 |
| | | Follow-up | 06/2018 | 27 | 20 | 3 | 4 | 0 | 100 | 1.3 | 0.9 | 3 | 1a | 4 | 4 | 2 | -2 | -2 | 1+ | 1+ | 1+ | 194 | 8 |
| 33 | REAL | Before-intervention | 06/2018 | 19 | 15 | 0 | 4 | 0 | - | 0.6 | 0.7 | 3 | 1a | 3 | 3 | 1 | -2 | -2 | 2 | 2 | 2 | - | - |
| | | | 06/2018 | - | - | - | - | - | 100 | - | - | - | - | - | - | - | - | - | - | - | - | 234 | 3 |
| | | After-intervention | 06/2018 | 25 | 19 | 2 | 4 | 0 | - | 0.6 | 0.9 | 3 | 1a | 4 | 3 | 1 | -2 | -1 | 1+ | 1 | 1 | - | 5 |
| | | Follow-up | 07/2018 | 25 | 17 | 4 | 4 | 0 | 100 | 0.4 | 0.4 | 3 | 1a | 3 | 3 | 2 | -2 | -1 | 2 | 1+ | 2 | 245 | 3 |
| 34 | CTRL | Before-intervention | 06/2018 | 21 | 19 | 0 | 2 | 0 | 95 | 0.1 | 0.1 | 3 | 1a | 2 | 3 | 0 | -2 | -2 | 0 | 2 | 2 | 136 | 6 |
| | | After-intervention | 06/2018 | 22 | 20 | 0 | 2 | 0 | - | 0.0 | 0.0 | - | - | - | - | - | - | - | 1 | 2 | 3 | - | - |
| | | | 06/2018 | - | - | - | - | - | - | - | - | 3 | 1a | 2 | 3 | 0 | -2 | -2 | - | - | - | - | 6 |
| | | Follow-up | 07/2018 | 23 | 21 | 0 | 2 | 0 | 90 | 0.1 | 0.1 | 3 | 1a | 3 | 4 | 0 | -2 | -2 | 0 | 2 | 2 | 160 | 6 |
| 35 | REAL | Before-intervention | 06/2018 | 22 | 18 | 0 | 4 | 0 | 100 | 0.7 | 0.5 | 3 | 1a | 3 | 3 | 2 | -2 | -2 | 1 | 1+ | 2 | 203 | 10 |
| | | After-intervention | 06/2018 | 25 | 22 | 0 | 3 | 0 | - | 0.8 | 0.4 | - | - | - | - | - | - | - | 1 | 1+ | 2 | - | 10 |
| | | | 06/2018 | - | - | - | - | - | - | - | - | 3 | 1a | 3 | 3 | 2 | -2 | -2 | - | - | - | - | - |
| | | Follow-up | 07/2018 | 26 | 23 | 0 | 3 | 0 | 100 | 0.6 | 0.4 | 3 | 1a | 3 | 3 | 2 | -2 | -2 | 2 | 2 | 2 | 193 | 8 |
| 36 | CTRL | Before-intervention | 06/2018 | 37 | 28 | 4 | 5 | 0 | - | 3.6 | 2.9 | 3 | 1a | 3 | 3 | 2 | -1 | -1 | 1+ | 1 | 1 | - | - |
| | | | 06/2018 | - | - | - | - | - | 100 | - | - | - | - | - | - | - | - | - | - | - | - | 156 | 26 |
| | | After-intervention | 07/2018 | - | - | - | - | - | - | - | - | 4 | 1a | 3 | 3 | 2 | -1 | -1 | - | - | - | - | 30 |
| | | | 07/2018 | 41 | 31 | 4 | 6 | 0 | - | 4.1 | 3.5 | - | - | - | - | - | - | - | 1+ | 1 | 1+ | - | - |
| | | Follow-up | 08/2018 | 41 | 31 | 5 | 5 | 0 | 100 | 3.9 | 3.0 | 3 | 1a | 3 | 3 | 2 | -1 | -1 | 1+ | 1+ | 2 | 182 | 27 |
| 37 | CTRL | Before-intervention | 07/2018 | 18 | 15 | 0 | 3 | 0 | 100 | 0.0 | 0.0 | 3 | 1a | 3 | 3 | 2 | -2 | -2 | 2 | 3 | 2 | 131 | 3 |
| | | After-intervention | 07/2018 | - | - | - | - | - | - | - | - | 3 | 1a | 3 | 3 | 2 | -1 | -2 | - | - | - | - | 3 |
| | | | 07/2018 | 18 | 15 | 0 | 3 | 0 | - | 0.0 | 0.0 | - | - | - | - | - | - | - | 2 | 3 | 2 | - | - |
| | | Follow-up | 08/2018 | 18 | 15 | 0 | 3 | 0 | 100 | 0.0 | 0.0 | 3 | 1a | 3 | 3 | 1 | -1 | -2 | 3 | 4 | 1+ | 190 | 3 |
| 38 | REAL | Before-intervention | 06/2018 | 25 | 21 | 0 | 4 | 0 | - | 1.1 | 1.4 | 3 | 1a | 3 | 3 | 2 | -1 | -2 | 1+ | 2 | 3 | - | - |
| | | | 06/2018 | - | - | - | - | - | 95 | - | - | - | - | - | - | - | - | - | - | - | - | 156 | 10 |
| | | After-intervention | 07/2018 | 25 | 19 | 2 | 4 | 0 | - | 0.8 | 0.9 | - | - | - | - | - | - | - | 1 | 1+ | 2 | - | - |
| | | | 07/2018 | - | - | - | - | - | - | - | - | 3 | 1b | 3 | 4 | 2 | -1 | -2 | - | - | - | - | 9 |
| | | Follow-up | 08/2018 | 23 | 17 | 2 | 4 | 0 | 100 | 0.7 | 0.5 | 3 | 1a | 3 | 3 | 2 | -1 | -2 | 1 | 1+ | 2 | 218 | 9 |
| 39 | REAL | Before-intervention | 07/2018 | 14 | 13 | 0 | 1 | 0 | 100 | 0.4 | 0.4 | 2 | 1a | 4 | 3 | 0 | -2 | -2 | 2 | 1 | 1 | 174 | 3 |
| | | After-intervention | 08/2018 | 14 | 13 | 0 | 1 | 0 | - | 0.4 | 0.4 | 2 | 1a | 4 | 3 | 0 | -1 | -1 | 1+ | 1 | 1 | - | 4 |
| | | Follow-up | 08/2018 | 14 | 13 | 0 | 1 | 0 | 100 | 0.4 | 0.4 | 2 | 1a | 4 | 3 | 1 | 0 | -1 | 1 | 1 | 1 | 186 | 4 |
| 40 | CTRL | Before-intervention | 07/2018 | 11 | 10 | 0 | 1 | 0 | 95 | 0.1 | 0.1 | 2 | 1a | 2 | 3 | 2 | -2 | -2 | 2 | 3 | 1+ | 152 | 4 |
| | | After-intervention | 07/2018 | 11 | 10 | 0 | 1 | 0 | - | 0.1 | 0.1 | 2 | 1a | 2 | 3 | 2 | -2 | -2 | 2 | 2 | 2 | - | 4 |
| | | Follow-up | 08/2018 | 15 | 14 | 0 | 1 | 0 | 100 | 0.1 | 0.1 | 2 | 1a | 3 | 3 | 2 | -2 | -2 | 2 | 2 | 1+ | 163 | 4 |

*Safety Analysis*

**Table S19** Overall adverse event incidence indexed by system organ class and preferred term

| Section | | REAL group | CTRL group |
|---|---|---|---|
| | | case (%) | case (%) |
| Number of cases | | 19 | 21 |
| Total adverse events | | 7 | 4 |
| Gastrointestinal disorder | | 2 | 1 |
| | Diarrhea | 1 (5.3) | 0 (0.0) |
| | Eructation | 1 (5.3) | 0 (0.0) |
| | Stomatitis | 0 (0.0) | 1 (4.8) |
| General disorders and administration | | 3 | 1 |
| | Dermatitis at the site of medical device use | 1 (5.3) | 1 (4.8) |
| | Irritation at the site of application of medical devices | 1 (5.3) | 0 (0.0) |
| | Pain at the site of application of medical devices | 1 (5.3) | 0 (0.0) |
| Infections and infestations | | 0 | 1 |
| | Nasopharyngitis | 0 (0.0) | 1 (4.8) |
| Musculoskeletal and connective tissue disorders | | 1 | 1 |
| | Arthritis | 1 (5.3) | 1 |
| | Myalgia | 0 (0.0) | 1 (4.8) |
| Skin and subcutaneous tissue disorders | | 1 | 0 |
| | Rash | 1 (5.3) | 0 (0.0) |
